# Vasomechanic treatment increases efflux of human brain proteins

**DOI:** 10.64898/2026.01.22.26344377

**Authors:** Johanna Tuunanen, Katariina Hautamäki, Tommi Väyrynen, Matti Järvelä, Vesa Korhonen, Niko Huotari, Mika Kaakinen, Janne Kananen, Heta Helakari, Lauri Raitamaa, Jari Jukkola, Sanna-Kaisa Herukka, Katariina Lauren, Ulla Salmi, Lauri Eklund, Maiken Nedergaard, Vesa Kiviniemi

## Abstract

An age-related decline in vasodilation mediated by neurovascular nitric oxide (NO) and vasoconstriction driven by the Piezo1 receptor precede the aggregation of soluble brain proteins such as amyloid-β (Aβ), which then causes further disruption of brain solute homeostasis, ultimately leading to neurodegeneration in Alzheimer’s disease. Preclinical studies show that restoring these vascular functions increases neurofluidic efflux and improves cognitive outcomes. Here, we tested effects of sublingual NO and/or Piezo1 receptor-targeted mechanotransductive whole-body vibrations (WBV_p_) in healthy adults (n = 29) on brain fluid dynamics and CNS-to-blood protein efflux using multimodal neuroimaging and blood biomarker analysis. The combined vasomechanic interventions (NO+WBV_p_) produced a synergistic enhancement of brain fluid transport and markedly increased the efflux of soluble brain-derived proteins (Aβ_40&42_, glial fibrillary acidic protein) into the bloodstream. The effects increase with age and the magnitude of NO-induced hypotension. Importantly, the combined intervention was well-tolerated, with no severe adverse physiological responses. Results demonstrate that a simple, non-invasive vasomechanic intervention can transiently promote brain-to-blood protein clearance in humans, highlighting a potentially safe and accessible therapeutic avenue for neurodegenerative conditions characterized by impaired brain solute removal and protein aggregation.

## Introduction

The efficient removal of soluble metabolites and proteins from interstitial and cerebrospinal fluids (I/CSF) is essential for maintaining brain homeostasis. Impairment of brain clearance pathways has been linked to the earliest stages of neurodegenerative disease, preceding neuronal loss by years or decades (Nedergaard and Goldman, 2020). Experimental and clinical studies demonstrate impaired clearance of soluble amyloid-β (Aβ) and phosphorylated tau protein from the CSF leads to their accumulation within perivascular and interstitial compartments, promoting aggregation and accelerated neuropathology (Seppälä et al., 2012; Olsson et al., 2016; Peng et al., 2016; Greenberg et al., 2020; Nedergaard and Goldman, 2020; Chen et al., 2025). For example, a murine model of Alzheimer’s disease exhibits progressive perivascular Aβ deposition accompanied by loss of endothelial low-density lipoprotein receptor-related protein 1 (LRP1), a key transporter mediating Aβ efflux to blood (Chen et al., 2025). Elevated soluble Aβ concentrations within CSF further impede perivascular transport, creating a feed-forward cycle in which Aβ transport becomes increasingly impaired (Peng et al., 2016; Chen et al., 2025). Therapeutic strategies that can safely enhance CSF solute clearance therefore represent a promising approach to delaying or preventing the onset of neurodegenerative disease.

Recent findings suggest that CSF efflux can be modulated through physiological mechanisms that are both therapeutically targetable and monitorable in humans. Acute systemic vasodilation caused by hypotensive drugs such as nicardipine and sodium nitroprusside (SNP) has emerged as a potent enhancer of CSF solute efflux in rodents (Jukkola et al., 2024). Nitric oxide (NO) donors such as SNP increase movement of intracisternal tracers from the cranial cavity to blood, predominantly via cervical lymph nodes, an effect that is accompanied by improved downstream lymphatic function (Jukkola et al., 2024; Yoon et al., 2024). Local NO delivery to neck lymphatic plexuses similarly enhances efflux of intracranial tracers, demonstrating that modulation of vascular tone can directly augment CSF transport along g/lymphatic pathways (Yoon et al., 2024).

Mechanical stimulation of Piezo1, a mechanosensitive cation channel expressed ubiquitously in all tissues including vascular and lymphatic endothelia, presents a complementary route for modulating neurofluid flow (Jin et al., 2025). Recent work shows that even light mechanical stimulation activates Piezo1, increases endothelial NO synthase (eNOS) activity, and restores age-related deficits in cervical lymphatic efflux (Wang et al., 2016; Jin et al., 2025). Piezo1 activation also increases meningeal lymphatic drainage (Choi et al., 2024; Matrongolo et al., 2024) and improves learning and memory in mice (Chi et al., 2022).

Although pharmacological Piezo1 agonists are not yet clinically available, mechanical stimulation is an alternative noninvasive way to activate the receptor. Thus, low-frequency (≤ 40 Hz) whole-body vibration (WBV) improved cognition, brain activity (Regterschot et al., 2014; Boerema et al., 2018) and motor function (Mosabbir et al., 2020; Yu and Kwon, 2024) in humans, and reduced neuroinflammation (Chen et al., 2022; Oroszi et al., 2022), and Aβ burden (Martorell et al., 2019) in animal models. In humans, a 40 Hz vibration stimulus of the head modulated brain pulsations and CSF–brain coupling (Kong et al., 2025), pointing to a translational pathway for mechanotransductive stimulation.

Here, we investigated whether NO-mediated vasodilation and/or Piezo1-targeted mechanical stimulation (WBV_p_) could enhance CSF-to-blood solute transport in humans. We exploited the capacity of dark-fluid magnetic resonance imaging (MRI) to detect protein-rich fluid, as demonstrated in prior studies using dark-CSF sequences to assess CSF protein burden (Ringstad and Eide, 2020; Albayram et al., 2022), to characterize physiological changes in brain water and macromolecule distribution. We combined these measurements with quantification of blood levels of brain-derived proteins that are relevant to neurodegeneration, including Aβ_40_, Aβ_42_, and glial fibrillary acidic protein (GFAP). Our results revealed a synergistic vasomechanic effect of combined NO and WBV_p_ stimulation on human CSF clearance, with participant age- and molecular size-dependent patterns of protein efflux, thus uncovering a previously unrecognized physiological efflux pathway in the human brain.

## Results

In this human crossover trial (Fig. S1), healthy volunteers (N = 29, of which 15 females, mean age: 44 ± 10 years; range 27-61 years) underwent three interventions in randomized order and on separate days: NO-mediated vasodilation (NO), mechanotransductive whole-body vibration (WPV_p_), and the combination protocol (NO+WBV_p_). This design allowed us to isolate the distinct physiological and blood chemistry effects of each intervention. We also conducted intrathecal tracer studies in mice to further investigate the underlying mechanisms.

### NO-mediated vasodilation combined to Piezo1-targeted whole-body vibration (NO+WBV_p_) enhances endogenous human brain to blood protein clearance

CSF tracer efflux has been shown to increase after NO exposure (Jukkola et al., 2024) and after Piezo1 activation in mice (Choi et al., 2024; Jin et al., 2025). We here tested the effects of NO-mediated vasodilation therapy combined with WBV_p_ (Fig. 1a) on brain protein efflux and physiology in 26 healthy participants (13 females; Fig. 1b).

**Figure 1.**
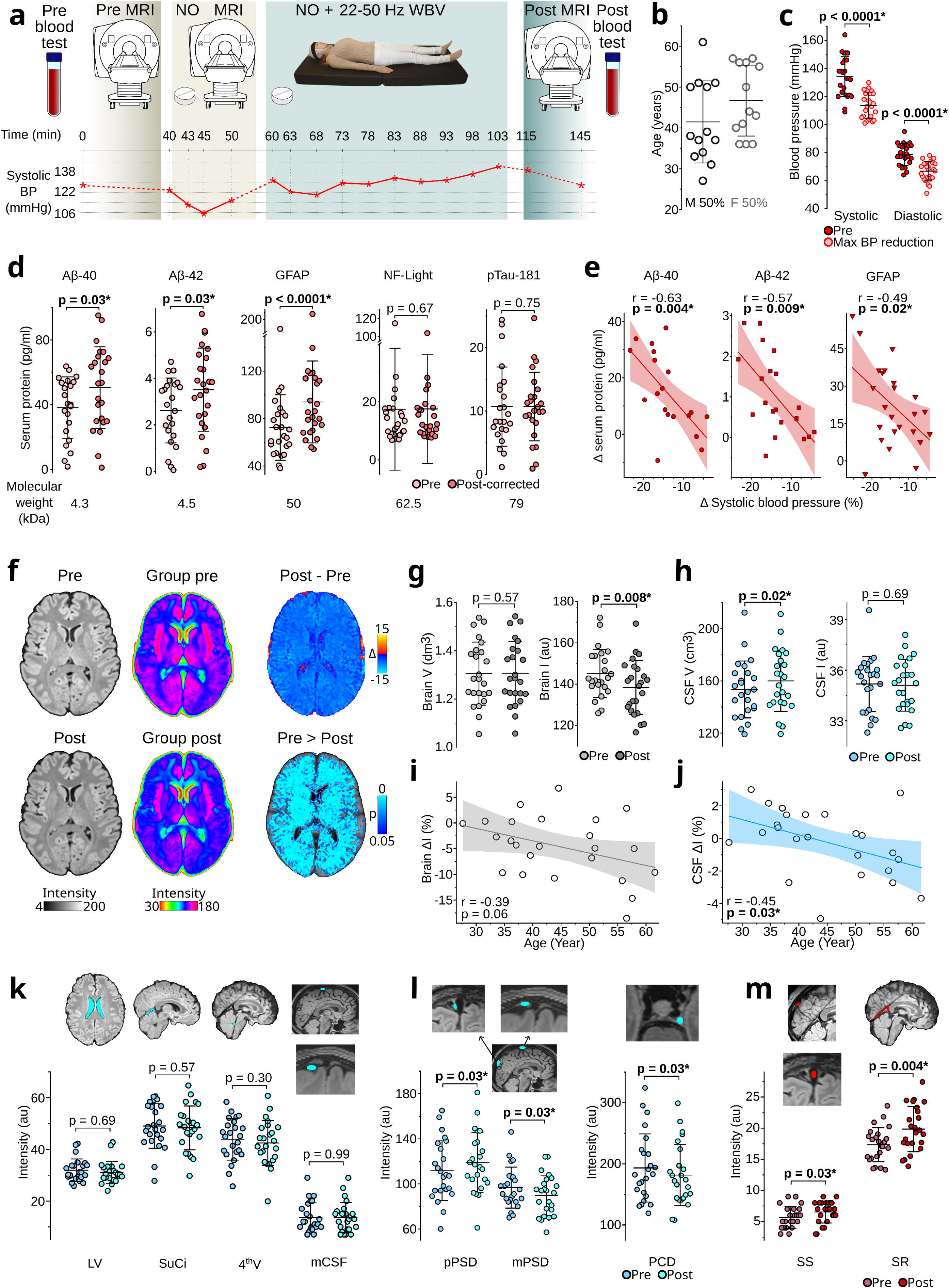
Human brain-to-blood protein clearance is enhanced by NO-mediated vasodilation combined with Piezo1-targeted whole-body vibration (NO+WBV_p_). **a,** Study design of the NO+WBV_p_ condition of the randomized crossover trial, and a chart of systolic blood pressure from a representative subject during the interventions. **b,** Age and sex distribution of participants (n = 26). **c,** The maximal reduction of blood pressure due to intervention. **d,** Venous blood analysis (Quanterix Simoa HD-X) before and after the NO+WBV_p_ intervention showing significantly increased beta-amyloid 40 & 42 (Aβ40, Aβ42) and glial fibrillary acidic protein (GFAP^#^), with unaltered levels of neurofilament-light (NF-Light) and phosphorylated tau-181 (pTau-181). Thus, low molecular weight (< 62 kDa) proteins increased in the systemic circulation, apparently in association with enhanced CSF efflux and e, in correlation with the individual extent of blood pressure reduction (Pearson’s correlation). **f,** T2 dark fluid (T2DF) images from a representative subject before and after NO+WBV_p_ intervention (Pre, Post), group-averaged color-coded T2DF images (Group pre, Group post), group averaged change map (Post – Pre), and statistical map indicating the declining brain signal intensity (two-sample paired t-test, family-wise-error-rate correction, p < 0.05) due to intervention (Pre > Post, n = 24). **g-h,** Segmented brain and CSF volumes and signal intensities from T2DF images showing that brain signal intensity decreased and CSF volume increased. **i-j,** Changes in brain and CSF T2DF signal intensities as functions of age (Pearson’s correlation), showing a strong age-dependence. Graphs indicate mean and 95% confidence intervals. **k,** ROI-analysis of T2DF images of lateral ventricles (LV^#^), superior cisterna (SuCi), 4^th^ ventricle (4^th^V), and middle parasagittal CSF space (pCSF^#^) showed no changes with intervention. l, In the CSF-filled spaces of the middle vertex parasagittal dura (mPSD) and para-cavernous dura (PCD), signal intensity decreased significantly, but increased in the posterior part parasagittal CSF space (PCSFs). **m,** ROI-analysis from posterior sinus sagittalis (SS^#^) and sinus rectus (SR) showed significantly increased T2DF signal intensity. All scatter plots show means, with whiskers extending to one standard deviation. Comparisons were performed using two-sided paired t-tests, with p-values adjusted for multiple comparisons using the Benjamini–Hochberg false discovery rate (p < 0.05) correction, unless otherwise indicated. ^#^Wilcoxon Signed Rank test was used for comparison.

Before treatment, systolic/diastolic blood pressure measured from the left brachium was 134 ± 14 / 79 ± 8 mmHg. During NO+WPV_p_ intervention, the pressure fell to a nadir of 114 ± 9 / 67 ± 7 mmHg (p < 0.0001, Fig. 1c, S2), corresponding to 14.9 ± 6.3 % (systolic) and 14.8 ± 9.0 % (diastolic) reductions.

To assess protein CNS-to-blood protein efflux, we collected venous blood samples before and after the interventions. Using the Quanterix Simoa HD-X platform to analyze serum (and plasma; Fig. S3b) concentrations, we found that the combined intervention significantly increased the low molecular weight (< 62 kDa) brain-derived proteins; Aβ_40_ (pre 38.2 ± 19.0 pg/ml, post hemodilution-corrected 50.5 ± 25.2 pg/ml p = 0.03), Aβ_42_ (pre 2.6 ± 1.4 pg/ml, post-corrected 3.5 ± 1.8 pg/ml p = 0.03), and GFAP (pre 65.2 (56.7 to 83.5) pg/ml (median (interquartile range)), post-corrected 85.1 (69.7 to 113.5) pg/ml, p < 0.0001) (Fig. 1d, S3a). In contrast, the larger neurofilament-light (p = 0.67) and phosphorylated tau-181 (p = 0.75) proteins were unaltered in serum, suggesting that molecular weight constrains CNS-to-blood efflux efficiency. Moreover, the magnitude of NO-mediated blood pressure reduction during NO+WBV_p_ treatment correlated with the increased serum concentrations of Aβ_40_, Aβ_42_, and GFAP (Fig. 1e).

S100 calcium-binding protein B (S100B, pre: 0.07 ± 0.10 µg/l, post 0.06 ± 0.04 µg/l, p = 0.4) and high-sensitivity C-reactive protein (hs-CRP, pre: 1.16 ± 1.26 mg/l, post 1.10 ± 1.12 mg/l, p = 0.08) showed no significant changes, suggesting a good margin of clinical safety. Furthermore, the measured GFAP levels were markedly lower than after severe traumatic brain injury (median: 1924 pg/ml) (Lei et al., 2015) or mild traumatic brain injury (mean: 1200 pg/ml) (Metting et al., 2012).

To assess treatment effects on cerebral fluid dynamics and water content, we compared 3D T2-weighted dark-fluid MRI (T2DF) anatomical high-resolution brain images acquired before and after the interventions; this approach is commonly used to evaluate brain edema, and more recently also for monitoring the presence of protein in parasagittal glymphatic CSF spaces (Ringstad and Eide, 2020; Albayram et al., 2022), c.f. Methods for details. Here, we used DF imaging parameters on a Siemens 3T scanner matching those in the latter study, which entailed protein calibration of the DF signal with respect to free water protein levels (Albayram et al., 2022).

Following treatment, whole-brain T2DF MRI signal intensity decreased by a mean of 4.6 ± 6.1 % (p = 0.008; Fig. 1f–g, 5c,g, S5), indicating a shift in the brain tissue water and/or macromolecule content in the brain, without any detectable change in brain tissue volume. In parallel, the volume of segmented extracerebral free dark CSF fluid increased by 6.5 ± 9.8 ml (p = 0.02; Fig. 1h, 5g). While the mean CSF signal intensity surrounding the brain did not change significantly, the individual pre vs. post intervention changes showed a significant inverse correlation with age (r = -0.45, p = 0.029; Fig. 1j); participants over than 45 years exhibited reduced CSF signal intensity, suggesting CSF water dilution, whereas younger individuals showed an increase in signal intensity more indicative of relatively increased protein efflux into CSF. Brain tissue signal intensity demonstrated a similar trend (r = -0.39, p = 0.062, Fig. 1i), with older participants showing greater post-treatment darkening. Representative brain tissue and CSF segmentation masks are provided in supplementary figure S20.

Encouraged by the anticorrelated whole-brain and CSF changes, we next examined the dark fluid MRI signal alterations within individually segmented tissue and fluid compartments to search for specific neurofluidic efflux sites. We performed individual region-of-interest (ROI) analyses to avoid spatial averaging artifacts introduced by MNI standard space registration, and keep our focus on structures previously implicated in glymphatic solute transport (Fig. 1k-m).

Within the parasagittal dura (PSD), the middle vertex PSD CSF-filled space (mPSD) showed a significant reduction in signal intensity (pre 97 ± 18 au, post 90 ± 18 au, p = 0.03, Fig. 1l), whereas the posterior parts PSD CSF space (pPSD), exhibited an increase (pre 112 ± 27, post 119 ± 27 au, p = 0.03, Fig. 1l). Signal intensity also decreased in the CSF-filled space in the para-cavernous dura (PCD, pre 193 ± 56 au, post 182 ± 50 au, p = 0.03, Fig. 1l).

In contrast, the T2 dark fluid MR signal increased within the venous sinuses (Fig. 1m), indicating increased protein level (Albayram et al., 2022); the intraluminal T2DF signal increased from 5.5 (4.0 to 7.0) au to 7.0 (5.5 to 8.0) au, (median (IQR)) p = 0.03) within the posterior superior sagittal sinus and from 17.3 ± 2.7 au to 19.9 ± 3.6 au, p = 0.004) in the sinus rectus. There was no significant change in the medial vertex segment of the sagittal sinus (pre 6.5 ± 2.0 au, post 6.9 ± 2.6 au, p = 0.67, Fig. S6b). Similarly, signal intensity was unaltered in CSF areas such as the lateral ventricles, superior cerebellar cisterna, fourth ventricles, or subarachnoid CSF space beneath the PSD, nor were there any changes in midline cervical lymph (pre 127.4 ± 20.4 au, post 127.3 ± 19.9 au, p = 0.99, Fig S6a). In summary, the T2 dark-fluid MRI results indicated that the combined vasomechanic treatment reduced the concentration of macromolecule-bound brain water molecules, suggesting an age-dependent increased clearance directly into the posterior and deep venous blood circulation, without concomitant brain volume loss.

Our supporting mice experiments with 40 Hz vibration and NO-donor SNP infusion demonstrated reduced accumulation of infused fluorescent FITC dextran (40 kDa) tracer in dorsal dura mater compared to SNP infusion alone, indicating that 40 Hz vibration reduces the concentration of macromolecules in dura (Fig. S22a).

### Vasomechanic NO+WBV_p_ effects on brain physiology

To assess how the NO+WBV_p_ intervention alters intracranial physiology and the pulsatile driving of brain hydrodynamics, we acquired multimodal whole-brain magnetic resonance encephalography (MREG, 10 Hz) recordings before and after intervention (Fig. 1a). Following the intervention, respiration-driven brain pulsation power increased in a focal region of the right temporal lobe (Fig. 2a-c, Supplementary Text1). Cardiovascular pulsation power (Järvelä et al., 2022; Tuunanen et al., 2024) was also elevated after the intervention (Fig. 2a-c, S7, Supplementary Text1). Fifteen minutes after the NO+WPV_p_ intervention, very-low frequency (VLF, 0.008 - 0.08 Hz) and low-frequency (LF, 0.08 - 0.15 Hz) vasomotor waves were unchanged compared with baseline (Fig 2a-b).

**Figure 2.**
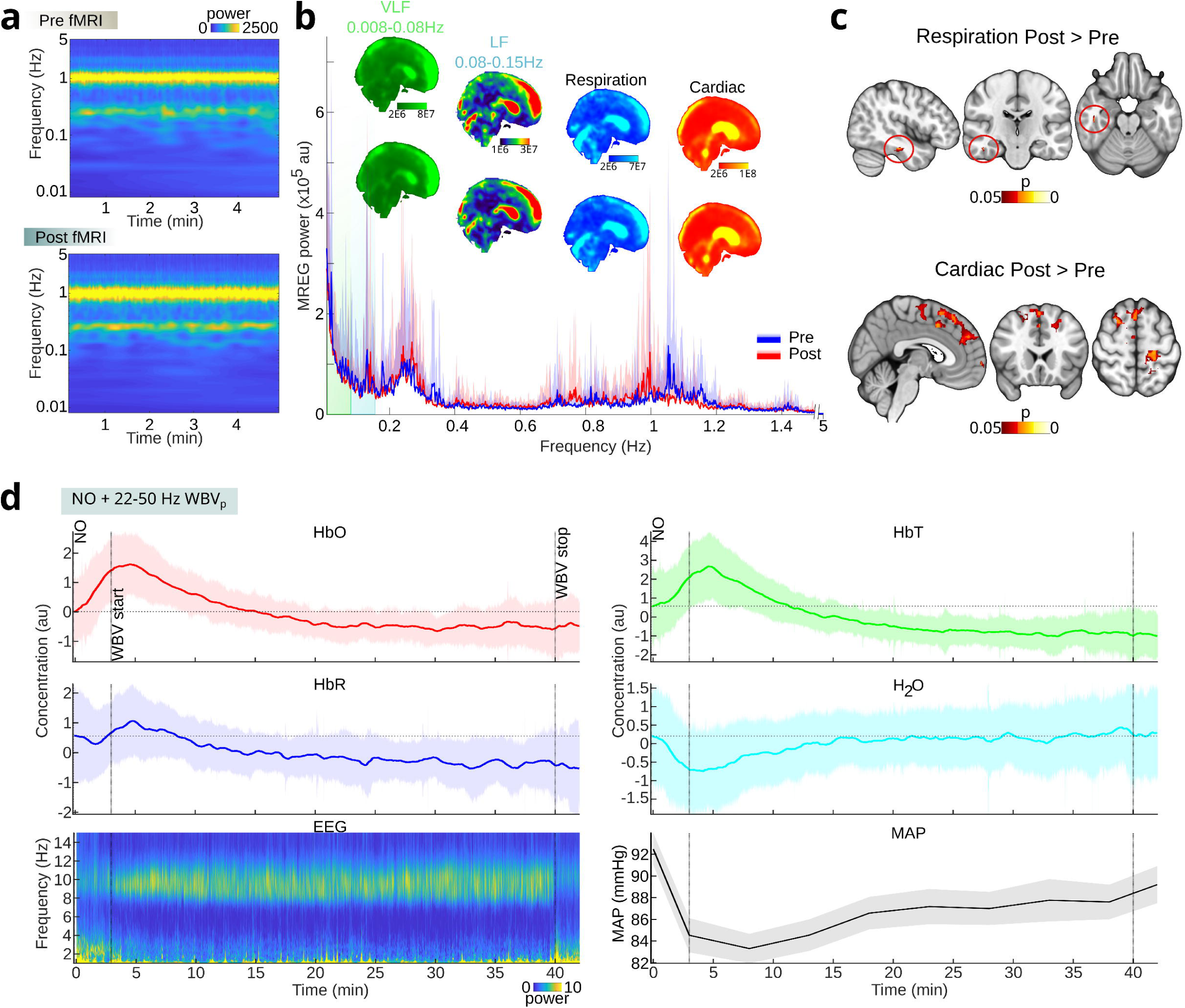
Combined vasomechanic NO+WBV_p_ intervention effects on brain physiology. **a-b**, Group average spectrogram and global power spectrum (mean+sd) with group mean power maps of very-low (VLF), low (LF), respiration, and cardiac frequencies from brain-wide ultrafast 10 Hz fMRI (MREG) scans (n = 23) taken before (Pre) and after (Post) NO+WBV_p_ intervention. **c**, Statistical maps of the voxelwise comparison between Pre- and Post-MREG show significantly increased pulsation powers in respiration and cardiac frequencies (two-sample paired t-test, family-wise-error-rate correction, p< 0.05). **d**, Electro- and hydrodynamic brain effect (n = 25) with near-infrared spectroscopy (fNIRS) measured at the forehead, and spectrogram of electroencephalography (EEG) recordings with simultaneous blood pressure measurements (n = 25) during the intervention. Group average fNIRS signal (showed mean ± standard deviation in the plots) of oxygenated blood (HbO), deoxygenated blood (HbR), total hemoglobin (HbT), and brain surface water content (H_2_O) corresponded to blood pressure changes (MAP = mean arterial pressure; mean ± standard error mean).

Because WBV_p_ is not compatible with MR scanning, we quantified physiological responses during the vasomechanical stimulus outside the scanner using wearable systems: functional near-infrared spectroscopy (fNIRS) over the forehead to measure hemo- (HbR/HbO) and H_2_O hydrodynamic and direct current electroencephalography (dcEEG) to assess electrophysiological responses during WBV_p_ experiments.

At the onset of intervention, there was a rapid rise in oxygenated hemoglobin (HbO) along with a slower and smaller increase in deoxygenated hemoglobin (HbR), consistent with NO-mediated vasodilation (Fig. 2d, Fig. 3k) (Meek et al., 1998; Kozberg and Hillman, 2016). These hemodynamic effects coincided with reductions in mean arterial pressure (MAP) and brain water content (H_2_O). MAP did not recover to baseline during the intervention, whereas H_2_O gradually returned to baseline after 37 minutes (Fig 2d). The combined hemo-hydrodynamic profile matched simple NO-mediated vasodilation during the first ∼10 minutes (Fig. 3k). Thereafter, concurrent WBV_p_ effects (Fig. 2d) also seen in the WBV_p_-alone experiment (Fig 4g) began to counteract the NO-driven changes, reversing the hemodynamic and hydrodynamic trends (Fig. 2d, Fig. 3k, Fig. 4g). Notably, the combined treatment evoked an H_2_O response distinct from that with WBV_p_ alone: while WBV_p_-alone caused a continuous rise in H_2_O accompanied by increased MAP (Fig. 4g), the combined NO+WBV_p_ intervention produced only a transient decrease in H_2_O, with subsequent recovery. Overall, the combined intervention seemingly reflects a temporal summation of NO and vibration effects, with a characteristic difference in brain water dynamics (Fig. 2–4).

**Figure 3.**
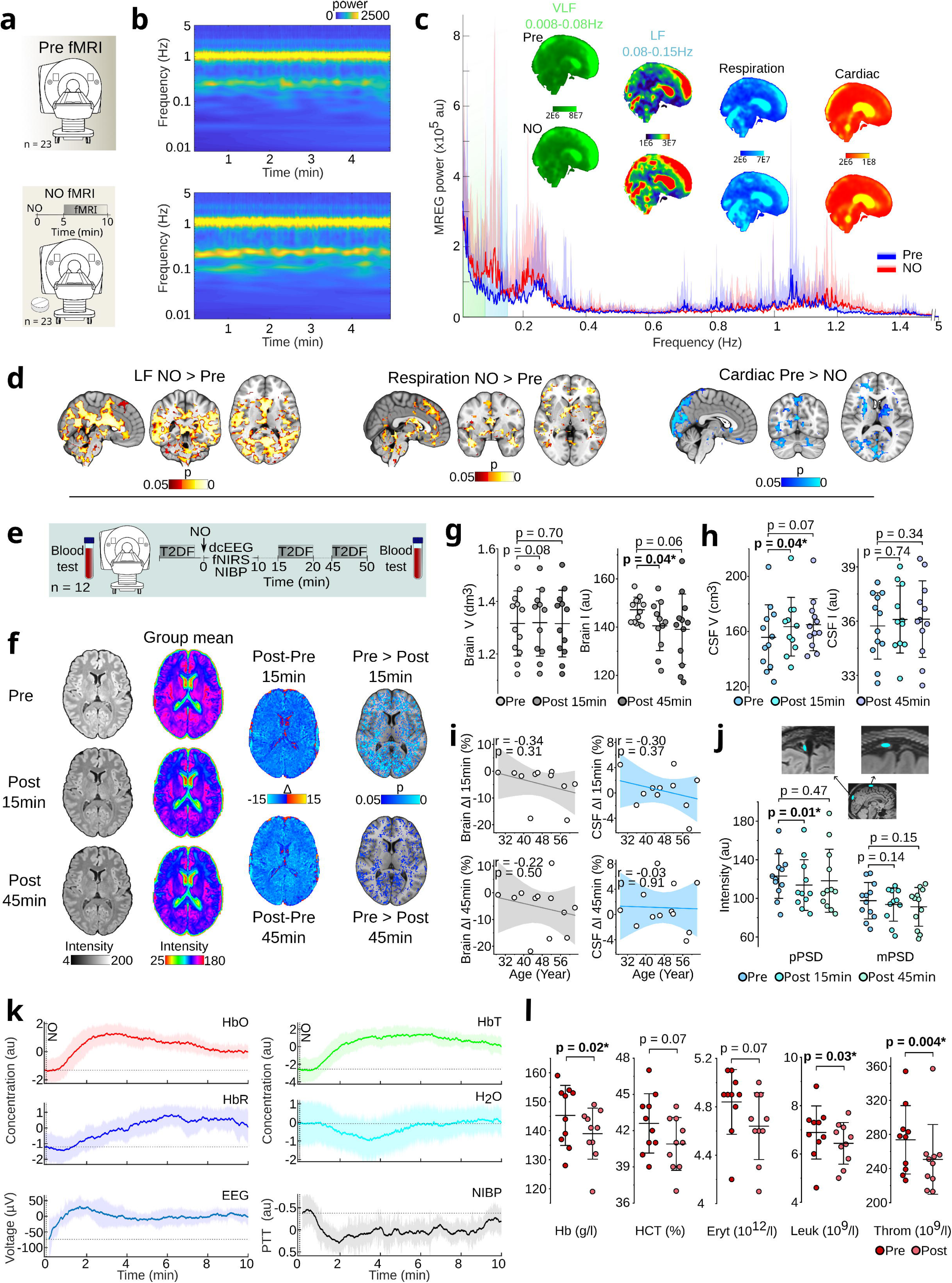
Isolated NO vasodilation (neuro)physiological effects. **a**, Subjects were scanned before taking 0.5 mg sublingual nitroglycerin. After blood pressure stabilization, they were scanned again for 5-minute with 10 Hz MREG. **b-c**, Group average spectrogram and global power spectrum with group mean power maps of very low frequency (VLF < 0.08 Hz), low frequency (LF 0.08-0.15 Hz), individual respiration, and cardiac frequencies from MREG measurements during NO-mediated vasodilation (n = 23). **d**, Statistical maps of the voxelwise comparison between baseline (Fig. 2) and during NO vasodilation MREG showed significantly increased pulsation powers in ∼0.1 Hz LF and respiration frequencies, whereas cardiac frequencies pulsation powers decreased (two-sample paired t-test, family-wise-error-rate correction (FWER), p < 0.05). **e**, Blood tests and water-sensitive T2-weighted dark fluid (T2DF) MRI images acquired before and after sublingual nitroglycerin from twelve subjects. **f**, Dark fluid images from a representative subject before and 15 min/45 min after nitroglycerin (Pre, Post 15min/45min), group-averaged color-coded T2DF images (Group pre, Group post 15/45), group averaged change map (Post 15/45 – Pre), and statistical maps indicating that brain signal intensity reduced significantly (two-sample paired t-test, FWER p < 0.05) due to NO vasodilation (Pre > Post 15/45). **g-h**, Segmented brain and CSF volumes and signal intensities from T2DF images showed that brain signal intensity decreased and CSF volume increased significantly. **i**, Changes in brain and CSF T2DF signal intensities showed no age dependency (Pearson’s correlation). Graphs depict mean and 95% confidence intervals. **j**, ROI-analysis of T2DF images of the posterior part of CSF-filled parasagittal dura space (pPSD) showed decreasing signal intensity, whereas there were no changes in the middle vertex parasagittal dura CSF space (mPSD). **k**, Electro- and hydrodynamic brain effects (n = 12) with near-infrared spectroscopy measured at the forehead, and electroencephalography (EEG) along with continuous simultaneous non-invasive blood pressure (NIBP, n = 12, PTT = pulse transit time) recordings during NO vasodilation. Group average signal (showed mean ± standard deviation in the plots) of oxygenated blood (HbO), deoxygenated blood (HbR), total hemoglobin (HbT), brain surface water content (H_2_O), and global EEG responded to estimated blood pressure changes (continuous NIBP; mean ± standard error mean). **l**, Blood test before and after NO-administration showed decreases in hemoglobin (Hb), hematocrit (HCT), erythrocytes (Eryt^#^) leukocytes (Leuk), and thrombocytes (Throm^#^), resulting in net 5.2 % dilution of the blood (n = 10). All scatter plots show means, with whiskers extending to one standard deviation. Comparisons were performed using two-sided paired t-tests, unless otherwise indicated. ^#^Wilcoxon Signed Rank test was used for comparison.

**Figure 4.**
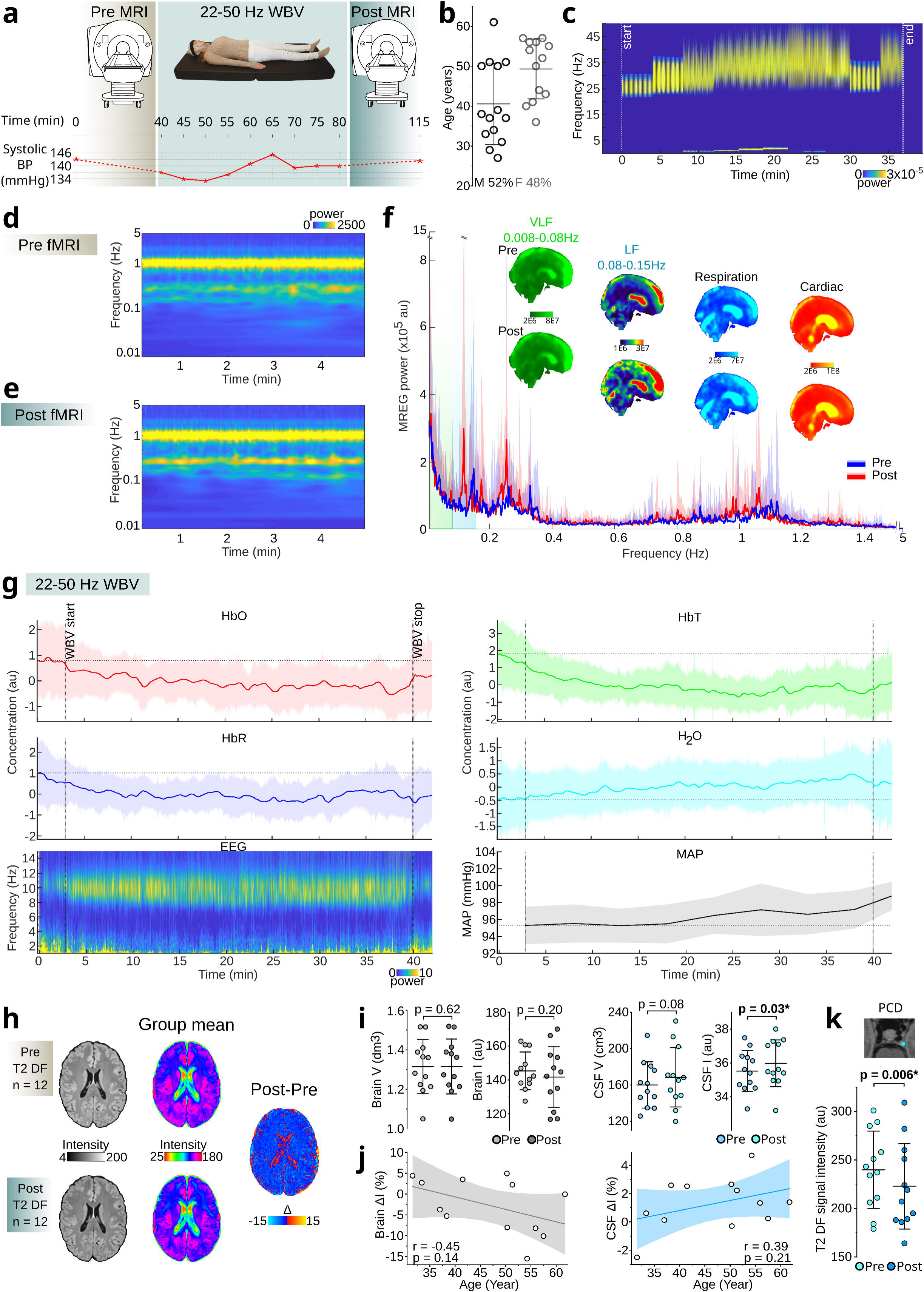
Piezo1-targeted whole-body vibration (WBV_p_) effect on brain physiology and water content. **a**, Study design of the WBV_p_ part of the randomized crossover trial, and a representative example of systolic blood pressure during the intervention. **b**, Age and sex distribution of participants (n = 27). **c**, Spectrogram of the accelerometer from the Piezo1-targeted (22 – 50 Hz) vibration element positioned under the head. **d-f**, Group average spectrogram and global power spectrum with group mean power maps of very low (VLF), low (LF), respiration, and cardiac frequencies from 10 Hz brain-wide fMRI (MREG) measurements (n = 25) before (Pre) and after (Post) WBV_p_ intervention showed no statistical changes of the voxelwise comparisons (two-sample paired t-test, family-wise-error-rate correction). **g**, Electro- and hydrodynamic brain effect with near-infrared spectroscopy (n = 22) measured at the forehead, and the spectrogram of electroencephalography (EEG, n = 25) with simultaneous blood pressure recordings (n = 27) during the intervention. Group average signal (showed mean ± standard deviation in the plots) of oxygenated blood (HbO), deoxygenated blood (HbR), total hemoglobin (HbT), and brain surface water content (H_2_O) responded to blood pressure changes (MAP = mean arterial pressure; mean ± standard error mean). **h**, T2 dark fluid (T2DF) images from a representative subject before and after intervention (Pre, Post), group-averaged color-coded T2DF images (Group pre, Group post, n = 12), and group-averaged change map (Post – Pre). There were no statistically significant changes in voxel intensity between the pre- and post-intervention, but **i,** segmented whole CSF signal intensity from the T2DF images showed a significant increase, along with a trend towards increasing volume. There were no changes in the brain volume, while a non-significant decrease in the brain intensity was observed. **j,** Change in brain and CSF T2DF signal intensities as functions of age (Pearson’s correlation). Graphs indicate mean and 95% confidence intervals. **k**, Para-cavernous dura CSF filled space (PCD) T2DF signal intensity decreased significantly with WBV_p_. Comparisons were performed using two-sided paired t-tests.

During WBV_p_, dcEEG showed increased alpha power (8–12 Hz), likely reflecting eye closure and relaxed drowsiness, as participants kept their eyes closed for ∼45% of the period with vibration. Low-frequency activity (< 4 Hz) decreased during vibration and re-emerged immediately afterward.

### NO vasodilation (neuro)physiological effects

To quantify intracranial physiological responses to acute NO vasodilation, we acquired 10 Hz MREG during sublingual NO administration in twenty-three subjects (Fig. 3a). NO produced arterial vasodilation accompanied by systemic blood pressure reduction in both MAP and NIBP measurements (Fig. 2d, 3k, S2). This vasodilation increased the power and reduced the frequency of the brain vasomotor wave LF peak (Fig. 3b-d, S10). VLF power remained unchanged, despite the expectation that ubiquitous NO vasodilation could suppress vasomotion. NO also increased the respiratory (Fig. 3b-d, S11) and decreased cardiovascular (Fig. 3b-d, S12) brain pulsation power.

To assess acute NO-mediated vasodilation changes in greater detail, we performed T2DF MRI, dcEEG, fNIRS and continuous non-invasive blood pressure (NIBP) recordings in twelve participants (mean age: 46 ± 10 years; range 27-61 years, 6 females) on a different day (Fig. 3e).

The electro/hydrodynamic measures indicated that the NO vasodilation alone produced effects resembling the early part of the combined intervention. Whole-brain T2DF MRI signal intensity decreased by 4.8 ± 6.8 % at 15 min post-administration (p = 0.04, Fig. 3f-g, S8). Over the entire 45-minute measurement period, mean signal intensity continued to decline by 5.5 ± 9.1 %, showing a trend toward significance (p = 0.06, Fig. 3f-g, 5b, S9). Brain tissue volume remained unchanged (Fig. 3g), whereas CSF volume again had increased by 7.0 ± 9.7 ml at 15 minutes (p = 0.04) and 9.2 ± 15.6 ml at 45 minutes (p = 0.07, Fig. 3h, 5f).

Dark-fluid signal intensity in CSF did not change significantly (Fig. 3h), and neither the brain nor CSF signal intensity correlated with age after NO-mediated vasodilation (Fig. 3i). The posterior PSD CSF-filled space (Fig. 3j) displayed an acute decrease in signal intensity at 15 minutes (p = 0.01), suggesting a vasodilation-induced drop in the local macromolecule/water ratio. Further ROI analyses showed no significant changes in sagittal sinus, lymph nodes, or the CSF compartment (Fig. S13-S15).

NO induced hypotension (Fig. 3k) was accompanied by an immediate 117 ± 98 µV DC shift in the dcEEG, followed by a delayed rise in arterial HbO with a temporal profile closely resembling the NIBP and dcEEG curve outlines (Fig 3k). A slower, transient reduction in brain H_2_O content emerged, which was followed by a gradual venous HbR increase. At 15 & 45 min after NO-alone administration, T2DF MRI recapitulated the global brain signal reductions observed with the combined NO+WBV_p_ intervention (Fig. 3f-g).

Because NO administration induces hemodilution (Arend et al., 1994), we quantified dilution effects after the 0.5 mg sublingual dose (Fig. 1d). Basic blood count before and after NO showed decreases in hemoglobin (4.2 ± 4.7 %), hematocrit (3.8 ± 5.9 %), erythrocytes (4.0 ± 5.6 %), leukocytes (5.8 ± 7.8 %), and thrombocytes (8.3 ± 5.3 %), corresponding to an estimated 5.2 % blood-water dilution (Fig. 3l). We applied this correction factor to all blood protein analyses (Fig. 1d). We also confirmed that the NO-mediated vasodilation by SNP in anesthetized mice promoted a significant 40 kDa FITC dextran removal from CSF to blood (Fig. S22b).

### Piezo1-targeted whole-body vibration

Mechanical low-frequency Piezo1 activation enhances glymphatic solute flow (Lim et al., 2024).To isolate the physiological effects of Piezo1-targeted (modulated in the low-frequency range between 22 – 50 Hz, c.f. Fig. 4c) WBV_p_, twenty-seven participants (Fig. 4b) underwent measurements of brain pulsation power, water content, and hemodynamic and electrophysiological responses (Fig. 4a).

During WBV_p_, MAP showed an upward trend accompanied by an increase in brain H_2_O content and a decrease in total blood volume (HbT, arterial HbO and venous HbR) measured by fNIRS (Fig. 4g). Participants kept their eyes closed for ∼48 % of the 37 minutes vibration time and the EEG alpha power during the vibration-alone attained approximately 12 % higher power than in the combined NO+WBV_p_ intervention (Fig. 2d). There was also a reduction in low-frequency power (< 4 Hz), which was less pronounced than in the combined NO+WBV_p_.

T2DF MRI, which was available for twelve participants, showed a trend toward increased CSF volume (Fig. 4h-i, 5a), whereas brain tissue volume and intensity remained stable. Whole-brain CSF intensity increased significantly (p = 0.03, Fig. 4i, 5e), which may reflect an increase in small-molecule content within the cortical CSF spaces; ROI analysis, as in the earlier interventions (Fig. 1k-l), revealed no significant changes in the lateral ventricles, superior cisterna, fourth ventricles, or parasagittal CSF space (Fig. S16). Likewise, ROI analyses detected no significant changes in sinus, lymph nodes, or PSD (Fig. S17-S18).

Change in brain tissue T2DF signal intensity showed a non-significant correlation with age (r = -0.45, p = 0.14, Fig. 4j), with older participants tending to exhibit greater signal reduction (i.e., darker brain tissue). Likewise, CSF intensity tended to increase more in older individuals as well (Fig. 4j). There was a significant intensity decrease in the para-cavernous dura CSF-filled space (PCD, pre 239.8 ± 38.2 au, post 222.8 ± 42.1 au, p = 0.006, Fig. 4k). Although MAP rose during active WBV_p_ (Fig. 4g), follow-up blood pressure measured 30 minutes later in 15 subjects showed a modest decline relative to baseline (systolic 134 ± 14 → 131 ± 12 mmHg, p = 0.2; diastolic 82 ± 7 → 77 ± 7 mmHg, p = 0.06).

Across VLF, LF, respiratory or cardiovascular brain pulsation powers, there were no statistically significant voxelwise differences between pre- and post WBV_p_ scans (Fig. 4d-f).

### Treatment side effects

Mild adverse effects were reported by some participants following administration of the study drug, with four reporting a headache and three reporting nausea. In one case, the study was discontinued due to the severity of nausea. None of the participants reported discomfort associated with the any of the whole-body vibration procedure. There were no serious adverse events.

## Discussion

In this study, we tested the hypothesis that human brain solute and protein efflux into the systemic circulation could be enhanced through vasodilative NO, vasoconstrictive WBV_p_, or combined vasomechanic interventions that dynamically modulate cerebral hemodynamics and CSF transport in healthy volunteers. Following the combined intervention, we observed significant increases in the blood concentration of certain low molecular weight (<62 kDa) brain-derived proteins, to an extent proportional to the maximal systolic blood pressure drop. Mechanistically, the combined stimulus produced a synergistic and age-dependent reorganization of neurofluid dynamics, reflected as coordinated changes in global brain tissue, CSF, and posterior venous sinus signals on dark-fluid MRI, which tracked hydrodynamic signatures detected through simultaneous NIRS monitoring. NO alone induced the expected MAP reduction, cerebral hyperemia, blood-water dilution, and a slowing and amplification of vasomotor and respiratory pulsations, resulting in net neurofluidic expansion and decreased dark-fluid MRI signal intensity due to reduced water/macromolecule ratios. In contrast, whole-body low-frequency modulated vibration targeting Piezo1 receptors (WBV_p_) gradually increased systemic MAP, elevated brain tissue CSF-water signals, and reduced both arterial HbO and para-cavernous dural signals. Together, these findings indicate that combining pharmacological vasodilation with mechanical vasoconstrictive stimulation recruits complementary vascular and CSF transport mechanisms capable of enhancing human brain solute clearance, corroborating previous research findings.

### Effect of vasodilation on brain protein efflux

Endothelial deficiency of NO promotes AD pathology (Austin et al., 2013) and conversely, a large Taiwanese cohort study recently reported a reduced risk of dementia in individuals treated with nitric oxide (NO)-based vasodilators (Zhou et al., 2023), consistent with other clinical evidence that strict blood pressure control lowers the risk of transition from mild cognitive impairment to dementia (He et al., 2025; Reboussin et al., 2025). Mechanistically, NO-based vasodilation produces a drop in systemic blood pressure, which increases ICP and enhances brain tracer efflux (Jukkola et al., 2024). This pressure-dependent stimulation of solute clearance provides a plausible physiological mechanism for the potential preventative effect of NO therapy on neurodegeneration.

Soluble Aβ40 is thought to be most neurotoxic amyloid species in early Alzheimer’s disease, and is consequently a favored target for interventive treatment (Tolar et al., 2021). At the same time, increased CSF concentrations of Aβ_40&42_ directly impede glymphatic transport, producing a feed-forward deficit in solute clearance (Peng et al., 2016). Aging further exacerbates this deficit, through a reduction in endothelial LRP1-mediated Aβ transport and loss of AQP4 polarization at astrocytic endfeet, together leading to diminished Aβ removal to blood (Kress et al., 2014; Chen et al., 2025). Present findings indicate that the soluble Aβ species can be mobilized to exit human brain using a combined vasomechanic NO+WBV_p_ intervention. This suggests a feasible, non-invasive approach to counteract the age-related decline in brain protein efflux, which would be potentially protective against AD. However, given our findings of upper size limit of approximately 62 kDa, such interventions would need to be applied before proteins such as Aβ or α-synuclein aggregate into larger oligomers or intravascular deposits.

Previously the physiological state most strongly linked to efficient glymphatic transport is sleep: CSF tracer efflux and removal of Aβ increase during natural sleep (Xie et al., 2013; Hablitz et al., 2020), while sleep loss in humans reduces Aβ clearance (Shokri-Kojori et al., 2018; Eide et al., 2022). During sleep, orexin-dependent reductions in central noradrenergic tone and blood pressure (Hauglund et al., 2025; Järvelä et al., 2025) induce robust vasomotor and respiratory pulsations to promote solute efflux (Fultz et al., 2019; Helakari et al., 2022; Tuunanen et al., 2024; Väyrynen et al., 2024; Hauglund et al., 2025). NO intervention appears to recapitulate key elements of this sleep-associated physiology; it lowers blood pressure and amplifies ∼ 0.1 Hz vasomotor brain waves and respiratory pulsations (Fig 3b-d) (Helakari et al., 2022, 2023), which likely together contribute to the observed enhancement of protein efflux into blood.

### Piezo1 receptor stimulus along with NO-mediated vasodilation induces synergistic brain water and protein efflux

Piezo1 channels in cerebrovascular endothelium are activated by mechanical forces, and counteract excessive arterial vasodilation by triggering vasoconstriction (Lim et al., 2024). Notably, overactivity to Piezo1 channels in vascular endothelium leads to learning and memory impairments in rodents, while an increase in astrocytic Piezo1 activity promotes the same cognitive function (Chi et al., 2022; Lim et al., 2024), highlighting their target specific relevance to neurocognition. Our WBV_p_ stimulation protocol evoked a gradual reduction in cerebral blood flow together with an increased MAP, consistent with a reactive vasoconstriction like that following Piezo1 activation (Fig. 4g).

Importantly, a precise low-frequency modulation of the vibration pattern is essential for effective Piezo1 activation in our approach, as the Piezo1 cation channels rapidly desensitize during prolonged stimulation (Lewis et al., 2017). Available pharmacological agonists of Piezo1, such as Yoda1, remain ineffective for brain applications because they fail to cross blood-brain-barrier (BBB) (Ikiz et al., 2024). Agonist drugs also cause tonic activation of endothelial Piezo1, which induces a long-lasting vasoconstriction that attenuates the important vasodilation responses coupled to neuronal activity (Lim et al., 2024).

Mechanotransductive vibration therapies has repeatedly been shown to improve learning, cognition, and motor function across multiple species (Regterschot et al., 2014; Boerema et al., 2018; Mosabbir et al., 2020; Chen et al., 2022; Oroszi et al., 2022; Wen et al., 2023; Yu and Kwon, 2024). Low-frequency vasomotor brain pulsations (Hauglund et al., 2025) driving hydrodynamics over BBB especially in sleep (Väyrynen et al., 2024), as well as mechanical stimulation of the cervical lymphatic plexus in mice increase CNS solute efflux (Jin et al., 2025). In contrast to pharmacological approaches, the WBV_p_ used in this study enables whole-body Piezo1 activation in a low-frequency oscillating pattern, which further augments brain tissue solute efflux (Fig. 4i). Importantly, the mechanical activation of Piezo1 in blood and lymphatic vessels further stimulates eNOS-driven NO production (Wang et al., 2016; Nonomura et al., 2018; Choi et al., 2024; Jin et al., 2025), offering a mechanistic basis for the synergistic enhancement of brain hydrodynamics and solute efflux seen with the present combined vasomechanic intervention (Fig. 1,5).

**Figure 5.**
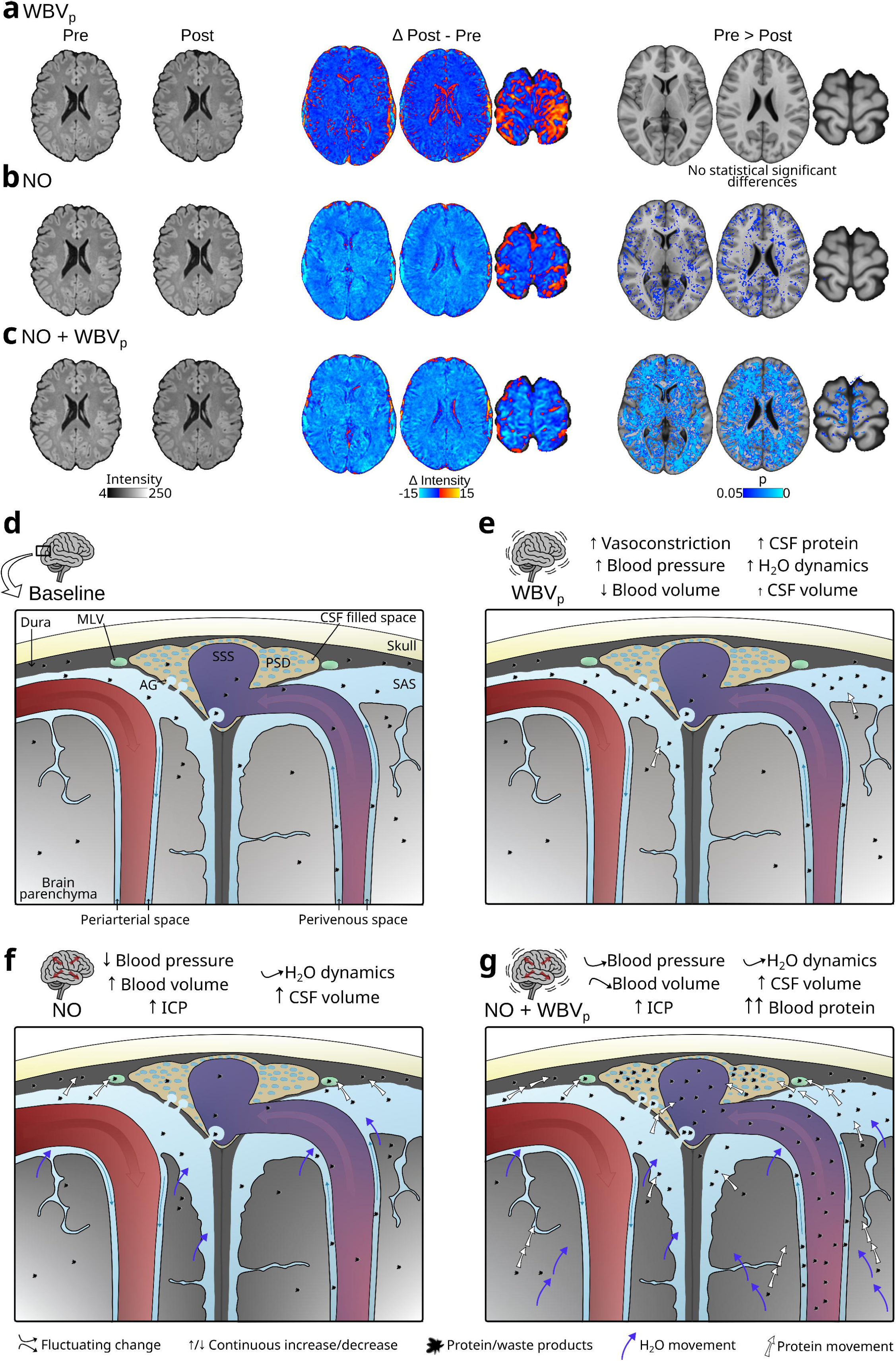
Brain-to-blood protein efflux is enhanced by vasomechanic NO+WBVp intervention. T2 dark fluid (T2DF) MR images from a representative subject before and after the intervention (Pre, Post), group averaged differential maps (Post – Pre, n = 12), and statistical difference maps (Pre > Post, n = 12, two-sample paired t-test, family-wise-error-rate correction p< 0.05) of **a.** WBV_p_, **b.** NO and **c.** combined NO+WBV_p_ interventions, indicating that the combined intervention had a synergistic effect in reducing brain signal intensity. In NO and NO+WBV_p_ interventions, mean and difference maps were calculated from images of the same participants. **d-g.** Illustrative diagrams of baseline and the main effects on Piezo1-targeted whole-body vibration (WBV_p_), NO-mediated vasodilation (NO) and NO-mediated vasodilation combined to Piezo1-targeted whole-body vibration (NO+WBV_p_) interventions. CSF = cerebrospinal fluid, ICP = intracranial pressure (Jukkola et al., 2024), SAS = subarachnoid space, SSS = superior sagittal sinus, PSD = parasagittal dura, ML = meningeal lymphatic vessel, AG = arachnoid granulation.

Ageing introduces several convergent deficits that degrade brain fluid transport, including reduced eNOS activity, diminished AQP4 expression (Colas et al., 2006; Choi et al., 2024) and endothelial Piezo1 function (Jin et al., 2025), which together impair (g)lymphatic efflux functions (Da Mesquita et al., 2018). These changes help explain the age dependency observed in the present hydrodynamic responses (Fig 1i-j, S4). Whole-body vibrations may also boost NO production via eNOS activation, thereby promoting multiple other neuroprotective pathways as well (Lopez et al., 2018). Within the brain, Piezo1 activation mitigated Aβ-induced defects in microglial motility and phagocytosis and improved Alzheimer disease -related cognitive dysfunction (Jäntti et al., 2022).

Based on the observed CSF and deep para-cavernous dural T2DF MRI changes (Fig. 1l, 4k), we propose that WPVp affects not only cortical glymphatic pathways, but also the deeper brain efflux routes that are traditionally associated with dural and basal outflow systems (Ringstad et al., 2018; Bohr et al., 2022). Supporting this view, our supplementary mouse experiments confirmed that dura solute clearance is enhanced by WPVp intervention (Fig. S22).

### Hydrodynamic brain changes during the interventions

A recent study indicated that solute efflux from the human brain can occur partly through perineural routes, as seen in rodents (Falkenberg-Jensen et al., 2025), but prior studies also suggest that the more permeable paraspinal CSF pathways are major contributors to net efflux (Melin et al., 2023). In the present human study, the dominant signature of the intervention was a whole-brain darkening of T2DF MRI signal, extending into deep structures (Fig. 1f, 3f). In clinical imaging, one usually looks for increased T2DF signal in tissue edema and darkening of T2DF signal typically reflects a decline of tissue edema after a successful therapeutic intervention. In line with this, brain edema in mice has been shown to decline upon NO-mediated vasodilation (Terpolilli et al., 2016).

Here, darkening of healthy brain tissue after the vasomechanic interventions likely reflects both a net shift of brain water into the CSF compartment (Fig 1f-h, 3h) and dilution (Arend et al., 1994) of blood constituents (Fig 3l). The parallel rise in low molecular weight soluble brain proteins (< 62 kDa) in blood further confirms an increased brain solute efflux (Fig. 1d), as also seen in the mouse model (Fig. S22b). The measured T2DF tissue darkening could thus reflect both reduced dissolved protein content in I/CSF and reduced ^1^H water proton signal, giving rise to lesser MR signal, as protein and water molecule concentrations both contribute to T2DF MRI intensity (Albayram et al., 2022).

The extent of increased brain solute efflux in healthy humans closely followed the magnitude of the blood pressure drop and age (Fig. 1e, S4). The simultaneously acquired fNIRS signal confirmed that brain water content indeed tracked the blood pressure drop during NO vasodilation (Fig. 2d). The changes in brain arterial HbO and slower venous HbR shifts were counteracted by the brain surface H_2_O signal, mostly reflecting subarachnoid CSF, in keeping with the Monro-Kellie principle governing intracranial fluid compartment dynamics.

We next examined dark-fluid MRI intensity in anatomical regions implicated in solute clearance using a dark fluid (a.k.a. FLAIR)-based technique as applied by others to investigate glymphatic PSD interfaces (Ringstad et al., 2018; Ringstad and Eide, 2020; Albayram et al., 2022). Post-intervention, blood in the posterior venous sinuses and in adjacent PSD regions showed increased dark-fluid signal, consistent with enriched local protein concentration. Importantly, MREG analyses showed that brain pulsations driving the neurofluids had returned to baseline during the dark-fluid MR imaging, making it unlikely that T2 flow-related changes explain these signal shifts. In contrast, the more frontal medial sagittal sinus and parasagittal/para-cavernous dural regions displayed reduced dark-fluid signal, consistent with simultaneous water dilution with increased protein removal (Fig. 1, S4), and suggesting a hydrostatic gradient that favors posterior solute drainage in the supine position.

Taken together, convergent T2DF MRI and blood biomarker results indicate a direct clearance route for brain-derived proteins from PSD regions into posterior venous sinuses. We conclude that human brain solute efflux can be safely and significantly enhanced with combined NO-mediated vasodilation and Piezo1-targeted mechanotransductive stimulation, acting through complementary effects on BBB transport, vascular hydrodynamics, and perivenous pathways.

## Methods

All human data was collected between 11/2021 - 10/2023. The study was approved by National Committee on Medical Research Ethics (Tukija), Finnish Medicines Agency (Fimea) and Regional Ethics Committee of the Northern Ostrobothnia Hospital District, and is registered to the EU Clinical Trials Register (clinicaltrialregister.eu, EudraCT number 2021-000625-27). The study was carried out in accordance with Medical Device Regulation (EU 2017/745), European Commission Directives on Clinical Trials (2001/20/EC) and Good Clinical Practice (2005/28/EC), Finnish National Legislation on Clinical Trials on Medicinal Products (488/1999) and Medicines (395/1987), ethical guidelines of the Helsinki declaration and the General Data Protection Regulation (EU 2016/679).

### Participants

The study involved 29 healthy volunteers (mean age: 44 ± 10 years; 15 females, 52%). Participants were recruited through email advertisements distributed to the personnel mailing lists of the University of Oulu and Oulu University Hospital. Inclusion criteria required participants to be healthy volunteers not using regular medication, except contraceptives or thyroid hormone replacement therapy (thyroxine). Exclusion criteria were pregnancy or breastfeeding, migraine or recurrent headache, substance or alcohol abuse, smoking or using tobacco products, known allergy to glyceryl trinitrate or other nitrates, current use of erectile dysfunction medication (sildenafil, vardenafil, or tadalafil), or contraindications for magnetic resonance imaging (including metal foreign bodies, severe claustrophobia, or electronic implants). Prospective participants contacted researchers voluntarily, underwent an initial phone interview, and subsequently attended an enrollment visit. During the enrollment visit, participants received detailed information about the study, voluntarily signed informed consent, and underwent medical screening by a physician to confirm their eligibility and ensure adequate health status for study participation.

### Study design

Eligible participants completed two intervention visits in randomized order: (1) *NO-mediated vasodilation with whole-body vibration* (NO+WBV_p_) and (2) *Piezo1-targeted whole-body vibration* (WBV_p_). After the first visit, each participant crossed over to the alternate intervention, resulting in a randomized crossover design. A washout period of at least seven days separated the visits. The study flow is summarized in the CONSORT diagram (Fig. S1). To investigate the acute effect of *NO-mediated vasodilation* (NO) alone in more detail (i.e., without WBV_p_), 12 participants underwent an additional NO session. Methods for the mouse study are presented in Supplementary text 2.

### Study drug

Sublingual nitroglycerin (glyceryl trinitrate, GTN, 0.5 mg per tablet; Nitroglycerin Orifarm®, Orifarm Healthcare a/s) was used as the study drug to cause vasodilation. GTN is an organic nitrate vasodilator that undergoes bioactivation to release nitric oxide (NO), which stimulates soluble guanylate cyclase in vascular smooth muscle cells, increasing intracellular cyclic guanosine monophosphate levels and thereby inducing smooth muscle relaxation. This mechanism produces potent vasodilation in arteries, with a predominant effect on venous capacitance vessels and significant dilation of large arteries (including the coronary arteries). When administered sublingually, GTN is rapidly absorbed through the oral mucosa, achieving peak plasma concentrations in approximately 2–5 minutes. The drug’s onset of action is thus almost immediate, and its hemodynamic effects persist for about 30–60 minutes. In clinical practice, sublingual GTN is widely used for the acute relief of angina pectoris.

### Whole-body vibration equipment

The Piezo1-targeting (Lewis et al., 2017) of the direct whole-body mechanotransductive stimulation was achieved by specific modulation of the 22-50 Hz whole-body vibration. The stimulus intensity modulation was carried out such a way that the Piezo1 mechanoreceptors would not saturate as they do with continuous tonic stimulus, but rather maximal triggering stimulus responses to > 20 Hz stimuli would be engaged after a refractory period, as shown in Fig. 4c. The stimulus was delivered with a commercially available whole-body vibration device (Neurosonic Mobile Mattress Basic, Oy Neurosonic Finland ltd).

### Multimodal data acquisition and preprocessing

Multimodal data were acquired simultaneously using magnetic resonance encephalography (MREG), MRI-compatible functional near-infrared spectroscopy (fNIRS), continuous non-invasive blood pressure (NIBP), cuff-based blood pressure monitoring, and direct-current electroencephalography (dc-EEG). All modalities were synchronized using scanner trigger pulses. Data were collected during a resting-state protocol, with subjects awake and fixating their gaze on a cross displayed on a screen. All data were acquired in Oulu University Hospital, Finland, using a single scanner setup and in synchrony with a previously described multimodal imaging protocol (Korhonen et al., 2014).

Preprocess steps and analyses were performed using MATLAB (R2024b, The MathWorks, Natick, MA), Functional MRI of the Brain Software Library (FSL), version 6.0.7.16) (Jenkinson et al., 2012) and Analysis of Functional NeuroImages (AFNI; version 18.0.05) (Cox, 1996).

#### (f)MRI data acquisition and preprocessing

MRI data were acquired using a Siemens MAGNETOM Vida 3T scanner (Siemens Healthineers AG, Erlangen, Germany) equipped with a 64-channel head coil. High-resolution anatomical T2 dark fluid (T2DF) weighted images were obtained using a standard Siemens DF space imaging in the sagittal plane (repetition time (TR) = 5000 ms, echo time (TE) = 387 ms, inversion time (TI)= 1800 ms, field-of-view (FOV) = 240 mm, flip angle (FA) = 120°, voxel size (VS) = 0.45 × 0.45 × 0.9 mm^3^).

Dark Fluid (T2DF) MRI is a one of the most important clinical MRI sequences as it is sensitive to edematous brain tissue water, which is detected as high signal intensity. The high T2DF signal stems from increased amount of water-^1^H surrounding brain macromolecules such as proteins. These sequences use an inversion pulse applied at specific inversion time (in humans at 37 °C, TI = 1800 ms), which nullifies signal from free moving, unbound CSF water molecule in DF images, whereas water affected by spin interactions with macromolecules such as proteins have a shorter T1 time and therefore give higher signal. Once the excess tissue water from an acute tissue edema resolves, the high T2DF signal darkens back to normal levels. In our study the DF images were used to quantitate changes in the brain tissue water/protein ratio caused by the interventions targeting vascular and perivascular structures.

The DF MRI was recently shown to also be advantageous for high precision estimation of key glymphatic structures like the parasagittal dura (Ringstad and Eide, 2020). As shown in phantom experiment by Albayram et al. (2022), increasing levels of protein (or other macromolecules) within free moving distilled water increases the dark fluid signal intensity. In the free-moving CSF and blood that appear as dark fluids in T2DF MRI, an increase in the signal may reflect an increase in the amounts of macromolecules, like soluble proteins, within the dark fluid. However, T2 flow effects can also increase the DF signal in fast flowing narrow CSF spaces, such as the aqueducts and sulci, but we avoided such narrowed spaces in our 3D ROI analyses by choosing broader CSF areas. Darkening tissue DF signal reflects a reduced amount of brain tissue water ^1^H in association with macromolecules, i.e., either free water ^1^H giving the MR signal, or associations with macromolecules changing the T1 relaxation of the ^1^H reduce, or both.

For further analysis, we used the FSL Brain Extraction Tool (BET) with neck and bias field correction for brain extraction from T2DF images. To confirm optimal quality, the extracted brain images were visually inspected.

T2-weighted images were obtained using a Siemens space imaging in the sagittal plane (TR = 3200 ms, TE = 412 ms, FOV = 240 mm, FA = 120°, and VS = 0.94 × 0.94 × 0.9 mm^3^) for defining lymph nodes and CSF-filled spaces within the dura from T2DF images (Fig. S19). T1-weighted images were obtained using a 3D MPRAGE sequence (TR = 1900 ms, TE = 2.49 ms, inversion time = 900 ms, FOV = 240 mm, FA = 9°, and VS = 0.94 × 0.94 × 0.9 mm^3^) for registration and for defining lymph nodes from T2DF images (Fig. S19). FSL BET with neck and bias field correction was used to extract the brain from the T1 images for registration of functional data to Montreal Neurological Institute (MNI152) space.

An ultrafast magnetic resonance encephalography (MREG) sequence was used to capture physiological brain pulsations at a temporal resolution of 10 Hz. MREG consists of a 3D single-shot stack-of-spirals sequence that under-samples k-space, enabling rapid whole-brain coverage (Assländer et al., 2013). The scanning parameters were: repetition time (TR) = 100 ms, echo time (TE) = 36 ms, flip angle (FA) = 25°, field of view (FOV) = 192 mm, voxel size (VS) = 3 × 3 × 3 mm^3^, and scanning time 5 min. Images were reconstructed using L2-Tikhonov regularization with λ = 0.1, determined via the L-curve method, and using a MATLAB-based reconstruction tool provided by the sequence developers (Hugger et al., 2011).

To minimize T1-relaxation effects, the first 180 time points were removed from the MREG data. MREG data are preprocessed with the FSL pipeline using high-pass filtration with a cut-off frequency of 0.008 Hz, and FSL MCFLIRT for motion correction. Additionally, the motion artefacts were minimized by excluding data that exceeded the following parameters: mean relative motion > 0.06 mm, mean absolute motion > 0.4 mm or absolute motion in any timepoint > 2 mm. MREG data were spatially smoothed with a 5 mm full width and half maximum Gaussian kernel. Any remaining motion-produced spikes were removed with AFNI. Finally, the MREG data was registered into MNI152 standard space using FSL for further analysis.

#### EEG and physiological data acquisition and preprocessing

Electroencephalographic (EEG) signals were acquired using a 32-channel MR-compatible direct-current dcEEG (Brain Products GmbH, Gilching, Germany). DcEEG recordings were obtained with a BrainCap MR EEG cap (EasyCap GmbH, Herrsching, Germany), in which Ag/AgCl electrodes were positioned according to the international 10–20 system. An electrolyte gel (ABRALYT HiCl, EasyCap GmbH) was applied between the electrodes and the scalp to ensure stable contact and to minimize impedance, which was maintained below 10 kΩ for all electrodes. DcEEG signals were recorded during MRI scanning using Brain Vision Recorder (Brain Products GmbH), and during WBV_p_ stimulation using the NeuroOne system (Bittium Corporation, Oulu, Finland), with a sampling rate of 5 kHz in both settings. Prior to measurements, we performed a visual inspection of signal quality. During MRI, the acquired dcEEG data (during NO intervention, n = 12) were preprocessed with Brain Vision Analyzer (v.2.1, Brain Products) to remove gradient and ballistocardiographic artifacts through template subtraction (Allen et al., 1998). Bad channels were identified by computing with MATLAB the standard deviation and maximum amplitude of each channel’s high-pass filtered (cutoff frequency 0.1 Hz) time series; channels with both SD > 2 and maximum amplitude > 1000 μV were excluded (0-1 channels per recording, 0.8 % overall). Filtered data were used only for identification of bad data to retain the dc-level. Because the polarity of DC shifts in EEG differs between individuals (Vanhatalo et al., 2003), we inverted the raw signals (multiplied by -1) so that slow shifts appear as upward (positive) deflections (0-31, 49.9 %). After that, data were downsampled to 500 Hz and a linear/quadratic trend was removed with MATLAB.

For dcEEG data acquired during the WBV_p_ mechanical stimulus, bad channels were identified by computing the standard deviation and maximum amplitude of each channel’s band-pass filtered (0.1 to 22 Hz) time series; channels with both SD > 2 and maximum amplitude > 1000 μV were excluded (NO+WBV_p_ intervention: 0-4 channels per recording, 3.2 % overall; WBV_p_ intervention: 0-3 channels per recording, 3.1 % overall). Next, the raw data were preprocessed in MATLAB using low-pass filtering with a cut-off frequency of 22 Hz to remove the vibration artefact.

Physiological signals were recorded in synchrony with fMRI (Korhonen et al., 2014), including respiration with respiratory belt and heart rate with fingertip photoplethysmography. Individual cardiorespiratory frequencies were calculated in MATLAB. Cuff-based blood pressure (systolic/diastolic) was recorded with an anesthesia monitor (GE Datex-Ohmeda Aestive 5).

#### (f)NIRS data acquisition and preprocessing

FNIRS data were collected using an MRI-compatible fiber-optic system (Korhonen et al., 2014) at a sampling rate of 800 Hz. High-power LEDs generated monochromatic light at wavelengths of 690, 830, and 980 nm. FNIRS optodes were attached to each subject’s forehead (right and left) at the source-detector distance of 3 cm to reach the brain cortex (Myllylä et al., 2018). Raw measured signals were processed to quantify with MATLAB the concentrations of oxygenated brain blood (HbO), deoxygenated brain blood (HbR), and cerebral CSF water below the skull (H_2_O) using a modification of the Beer-Lambert law (Myllylä et al., 2018). The total hemoglobin (HbT) signal was calculated as a sum of HbO and HbR signals. Afterwards, the group mean (and standard deviation) was calculated for the normalized right and left signals.

#### NIBP data acquisition and preprocessing

Continuous non-invasive blood pressure (NIBP) was measured using a fibre-optic accelerometer system optimized for MRI compatibility with sampling rate of 1 kHz (Korhonen et al., 2014). Two sensors were attached on the sternum over the aortic valve and carotid artery to sense skin movements caused by cardiovascular pulses. Pulse transit times were calculated between the sensors for each recording.

### Multimodal data analysis

For the MREG recordings, we used complex Morlet wavelets in wavelet convolution to calculate the time-frequency spectrogram (Väyrynen et al., 2024) with a fixed number of wavelet cycles (*N*=8) and logarithmic frequency ranging from 0.01 to 5 Hz with 50 equal steps. To reduce computational cost, we randomly selected 5% (3405 voxels) of brain voxels.

Global fast Fourier transform (FFT) power density maps were calculated using AFNI for the full band MREG data, followed by application of the FSL to calculate the average FFT spectrum of all brain voxels. The sums of VLF, LF, individual respiratory, and cardiac powers were calculated with FSL and AFNI. The VLF range of 0.008–0.08 Hz was chosen to obtain the maximum possible coverage of the very low frequencies, without overlapping with LF and respiratory frequencies. The 0.08–0.15 Hz band was selected to capture low-frequency ∼0.1 Hz vasomotor waves without overlapping with respiration.

Respiratory and cardiac ranges and peak frequencies were obtained from individual FFT spectrums without harmonics or overlapping with other physiological frequency bands. Frequencies were additionally double-checked from the scanner’s physiological signals (respiration belt and photoplethysmogram). Paired statistical tests excluded datasets exhibiting physiological frequency overlap, specifically for cases in which the respiratory frequency range overlapped into the VLF band. The resulting sample sizes for each comparison are reported in supplementary figure S21.

#### Anatomical T2 dark-fluid MRI data analysis

1. Whole-brain voxelwise comparison: T2-DF brain-extracted and bias-field corrected volumes were compared between pre- and post-intervention scans.
2. CSF/brain-tissue segmentation: coarse two-class segmentation masks (CSF vs brain tissue) were generated with FSL. Masks were refined using FSL with threshold values < 70 DF intensity (au) defined as CSF and > 70 defined as brain tissue. All masks were visually inspected. A representative example of the masks can be found in supplementary figure S20. Next, we calculated the mean intensity values and volumes with FSL.
3. Region of interest (ROI) analysis: ROI analysis was performed on T2DF images to obtain the individual mean intensity within each ROI before and after the interventions. ROIs were delineated using a diagnostic DICOM viewer and included the following: subarachnoid CSF beneath the PSD, middle/vertex PSD CSF-filled space, posterior PSD CSF-filled space, CSF-filled space in the paracavernous dura, posterior superior sagittal sinus, vertex/midline sagittal sinus, and superficial cervical lymph nodes. In addition, the following individual ROIs were drawn with the fsleyes tool: lateral ventricles, superior cisterna, fourth ventricle, and sinus rectus.

#### EEG data analysis

For EEG acquired during vibration, we used complex Morlet wavelets in wavelet convolution to calculate a time-frequency spectrogram (Väyrynen et al., 2024) with fixed number of wavelet cycles (*N*=8) and frequency range from 0.1 to 22 Hz in equal 220 steps for each EEG channel. We next calculated the group average EEG spectrogram, and then the corresponding alpha power (8 – 12 Hz). For EEG acquired during MRI, we calculated group means (and SD).

#### NIBP data analysis

PTT is inversely related to blood pressure, and can be used for cuffless blood pressure estimation (Hoshide et al., 2022). Generally, as blood pressure increases, the arterial walls become stiffer, leading to faster pulse wave velocity and shorter PTT. From each PTT data set, we calculated a moving average in MATLAB, and subsequently the group mean (and standard error of the mean).

### Blood test

Venous blood was drawn immediately before and after the NO+WBV_p_ intervention. EDTA-plasma and serum were separated and aliquoted into Sarstedt polypropylene tubes, frozen and stored at –70 °C, and shipped to the Eastern Finland Biomarker laboratory for analysis. Biomarkers were quantified on Quanterix’s Single Molecule Array (Simoa) HD-X Analyzer (Quanterix, Billerica, MA, USA) using the Neurology 4-PlexE assay (Aβ_40_, Aβ_42_, GFAP, NF-light) and the pTau181 Advantage V2.1 assay. For safety monitoring, the serum hs-CRP and S100B levels were measured by Nordlab (Oulu, Finland).

To evaluate the potential NO-induced hemodilution, a complete blood count (including hemoglobin, hematocrit, red and white blood cells and platelets) was obtained immediately before and after the NO intervention (n = 10). The analyses were performed by Nordlab (Oulu, Finland).

### Statistical analysis

Voxelwise comparisons between brain pulsation power maps and T2DF maps were performed by a two-sample paired *t*-test using a non-parametric threshold-free permutation test (5000 permutations) with FSL’s randomise (Smith and Nichols, 2009), and were corrected for the family-wise error rate (FWER) at a significance level of *p* < 0.05.

For the blood pressure data, segmented and ROI-based T2DF data, and blood results, we used the Shapiro-Wilk test to examine data normality. For normally distributed data, we applied a paired t-test, and for non-normally distributed data, the Wilcoxon signed rank test, using Origin Pro 2023b for the statistical analyses.

To control for the number of false positives among significant results of segmented and ROI analysis (n = 24) and blood results (n = 25) in the NO+WBV_p_ condition we used Benjamini-Hochberg false discovery rate (FDR) correction (Benjamini and Hochberg, 1995) with MATLAB.

Significant threshold for all tests was p < 0.05. All data were reported as meanL±Lstandard deviation (SD) unless otherwise indicated. Non-normally distributed data were reported as median (interquartile range, IQR). In Fig. 1e, outliers in the Pearson correlation analysis (> 2.5SD, 3 Ab40 & -42 and 1 GFAP) were masked before the final correlation analysis. The resulting sample sizes for each multimodal analysis are reported in Supplementary Fig. 22.

## Data availability

Sharing of individual-level MRI/fMRI data is governed by the Finnish Act on the Secondary Use of Health and Social Data and the European Union’s General Data Protection Regulation (GDPR), which restricts access when there is a risk of re-identification. Accordingly, data sharing is considered case-by-case upon reasonable request. Pseudonymized individual MRI/fMRI datasets may be shared with qualified academic collaborators after ethical review and data protection assessment, and subject to an approved data usage agreement. Due to these legal constraints, the original raw MRI/fMRI datasets cannot be deposited in a public repository. Requests should be directed to the corresponding author.

## Competing interests

The authors declare no competing interests.

## Acknowledgements

This study was funded by Research Counsil of Finland: TERVA grants 1-2, 275342, 338599, 314497, 335720 (V.Ki), 360508 (J.K), The EU Joint Programme – Neurodegenerative Disease Research 2022-120 (V.Ki), Jane & Aatos Erkko Foundation: 1, 210043 (V.Ki.), VTR grants from Oulu University Hospital (V.Ki, V.Ko, J.K). Instrumentarium Science Foundation Sr (T.V, J.T, M.J), The Finnish Medical Foundation (V.Ki, M.J), Finnish Brain Foundation (V.Ki, L.R, J.T), Pohjois-Suomen Terveydenhuollon tukisäätiö (H.H, V.Ko), Tauno Tönning Foundation (L.R), Juhani Aho Foundation for Medical Research (L.R, T.V), Uniogs/MRC Oulu DP-grant (H.H), Emil Aaltosen Säätiö (H.H, L.R, M.J). Finnish Cultural Foundation (M.J), Orion Research Foundation Sr (H.H, M.J, L.R, J.T.), Päivikki and Sakari Sohlberg Foundation (L.R), The Paulo Foundation (H.H, M.J), Thelma Mäkikyrö foundation (V.Ko). We thank CSC – IT Center for Science Ltd., Finland for providing high-performance computational resources used in this study. We further thank The Biomarker Laboratory of University of Eastern Finland for their collaboration. We thank Adj. Prof. Paul Cumming of Queensland University of Technology, Brisbane for comments on the manuscript.

## Notes

### Competing Interest Statement

The authors have declared no competing interest.

### Clinical Trial

EudraCT 2021-000625-27

### Author Declarations

Ethics committee/IRB of National Committee on Medical Research Ethics (Tukija) gave ethical approval for this work.

