## Supplementary information for "Vasomechanic treatment increases efflux of human brain proteins"

### Supporting information Text 1

#### Brain pulsation ranges from MREG

Individual respiratory and cardiac frequencies were extracted from the MREG periodogram using the MATLAB fast Fourier transformation function and double-checked for quality from the scanner respiration belt and photoplethysmogram data (Järvelä et al., 2022; Kananen et al., 2022; Tuunanen et al., 2024). From the signal frequency spectrum, respiratory and cardiac minima, maxima, and peak values were obtained and used to calculate individual cardiorespiratory frequency ranges.

#### Vasomechanic nitric oxide mediated vasodilation with Piezo1-targeted whole-body vibration (NO & WBVp)

Compared to baseline, the vasomechanic NO+WBVp intervention increased the spectral respiration range (Pre: 0.19 ± 0.05 Hz, Post: 0.21 ± 0.05 Hz, $p = 0.04$) but did not alter respiration rate (Pre and Post: 15 ± 4 b/min, $p = 0.97$). Heart rate also decreased (Pre: 62 ± 6 b/min, Post: 58 ± 8 b/min, $p = 0.001$), but the width of the cardiac range was not significantly unaffected (Pre: 0.25 ± 0.06 Hz, Post: 0.28 ± 0.05 Hz, $p = 0.051$).

#### Nitric oxide (NO) mediated vasodilation

NO-mediated vasodilation reduced respiratory rate (15 ± 4 b/min to 13 ± 4 b/min, $p = 0.007$) and increased heart rate (from 62 ± 6 b/min to 66 ± 9 b/min, $p < 0.0001$) compared to baseline. There were no significant effects on spectral respiration range (Pre: 0.19 ± 0.05 Hz, during NO 0.20 ± 0.06 Hz, $p = 0.363$) or cardiac range (Pre: 0.25 ± 0.06 Hz, during NO 0.27 ± 0.07 Hz, $p = 0.164$).

#### Piezo1 targeted whole-body vibration ($WBV_p$)

Compared to baseline, $WBV_p$ interventions did not markedly alter the spectral respiration range (Pre: 0.20 ± 0.06 Hz, Post: 0.20 ± 0.05 Hz, $p = 0.43$) and respiration rate remained stable (Pre 15 ± 4 vs. Post 15 ± 4 breaths/min, $p = 0.75$). Heart rate decreased slightly (61 ± 7 → 59 ± 8 beats/min, $p = 0.04$) and the spectral range of cardiac rate increased (Pre: 0.23 ± 0.04 Hz, Post: 0.25 ± 0.05 Hz, p = 0.02) significantly during the $WBV_p$ intervention.

### Supporting information Text 2

#### Methods for mice study

*Mouse study design*

2-3 months old C56 Bl6/N (Charles River) female mice were used in all experiments. Animals were housed in temperature- and humidity-controlled rooms with a 12 h light/12 h dark cycle. Teklad Global Rodent diet (Harlan Teklad, USA) and untreated tap water were offered *ad libitum*. Mice were anaesthetized with subcutaneous injection of ketamine (75 mg/kg) and xylazine (10 mg/kg) (KX), with supplementation of anaesthesia every 30 minutes during the entire experiment. Subcutaneous lidocaine (10 mg/kg) was administered to surgical sites 5 min before operations. All experimental procedures and animal care were in accordance with the Finnish and European legislation and were approved by the national Project Authorization Board (license numbers ESAVI/41363/2019 and ESAVI/2362/04.10.07/2017).

*Cisterna magna injections, combination and NO-mediated vasodilation with SNP treatment*

The femoral vein and cisterna magna (CM) cannulation and tracer infusion was done as described earlier (Jukkola et al. 2024). The femoral vein was exposed through an incision on skin and cannulated with a BTPE 50 polyethylene tubing filled with physiological saline supplemented with 500 IU/ml heparin. Next, BTPE-10 tubing, connected to a 30G dental needle, was inserted in the *cisterna magna* (CM) and fixed in place with superglue and dental acrylic. 4 µl (25 mg/ml) of 40 kDa of fluorescein isothiocyanate-dextran (Sigma, FD40S-250MG), was infused into the subarachnoid space at a rate of 2µl/min, driven by a microinjector (Legato130, KD Scientific). **Combination treatment:** After the CM-cannula was sealed with a cauterizer to prevent backflow, animals were placed onto a vibrating mattress on a supine position. A heating lamp was placed above the animal to keep their body temperature ~37ºC. Sodium nitroprusside (SNP) (Sigma, 10 µg/kg/min) was infused into the femoral vein for 15 min at a rate of 10 µl/min with or without 40 Hz vibration. Euthanasia was performed with an overdose of KX, followed transcardial perfusion with ice-cold 1x PBS supplemented with 500 IU/ml heparin. The dura mater from the dorsal part of the skull was collected, flash frozen in liquid nitrogen, and stored at -80 ºC. **NO-mediated vasodilation treatment:** 40 kDa FITC-dextran (FD40) injection into the CM was followed by blood sampling from the femoral vein 5 min later, with an i.v. infusion of SNP or saline (Control) starting 2 min after the first blood sampling and continuing for 40 min, after which the final blood sampling was performed. SNP (Sigma, PHR1423-1G) (5 µg/kg/min) was infused into the femoral vein at the rate of 5 µl/min for 40 minutes. The control group received 0.9% NaCl (saline) i.v. at a similar rate. Blood was collected and centrifuged at 1100 x g for 10 min and supernatant (serum) stored at -70 °C.

*Fluorescence measurements from serum and tissue*

The tissue samples and serum were thawed, weighed, and immersed in PBS and homogenized for 5 min at 50 Hz with a homogenizer (Qiagen, Tissue Lyzer LT) and stainless-steel beads. After centrifugation at 12,400 g for 1 hour at room temperature, the supernatant was collected and diluted in PBS. All diluted samples were assayed in triplicate using a 96-well plate. Serial dilutions of 40/4 kDa FITC dextran was used for making a standard curve. Fluorescence of FITC-dextran was measured with fluorometer ((PerkinElmer, VICTOR3V 1420 Multilabel counter) with excitation/emission at 485nm/535nm. Serum and dura mater samples without tracer were used as negative controls.

### Supporting figures

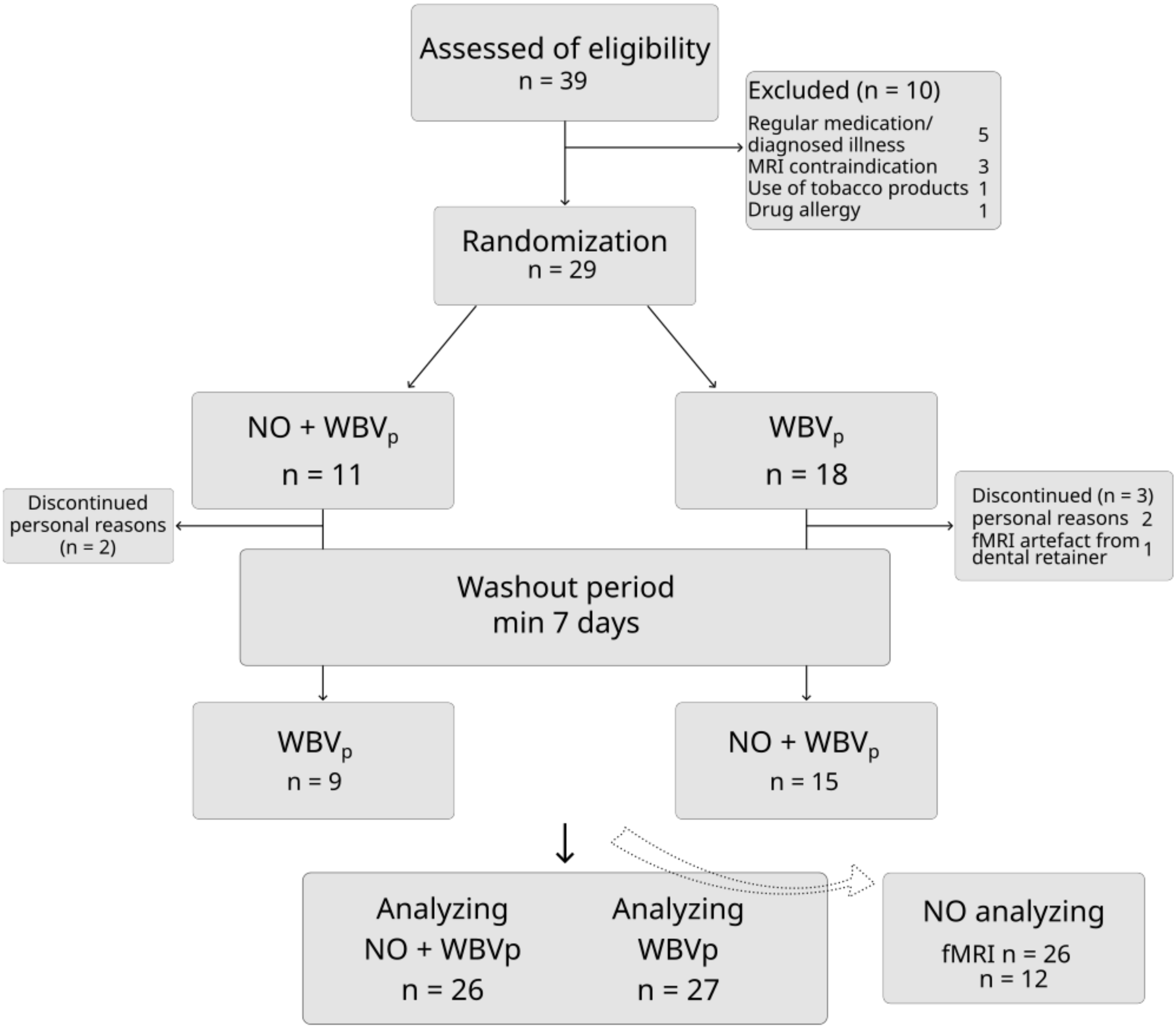


Figure S1. CONSORT (Consolidated Standards of Reporting Trials) flow diagram of randomized crossover design and exclusion of subjects. Twenty-nine subjects participated in a combined NO-mediated vasodilation with whole-body vibration (NO+WBVp, n = 26) intervention and whole-body vibration (WBVp, n = 27) intervention in randomized order on different days. Twelve of these participants also engaged in a NO-mediated vasodilation (NO) intervention, on a different experimental day.

**Vasomechanic nitric oxide-mediated vasodilation with Piezo-1 targeted whole body vibration (NO & WBVp)**

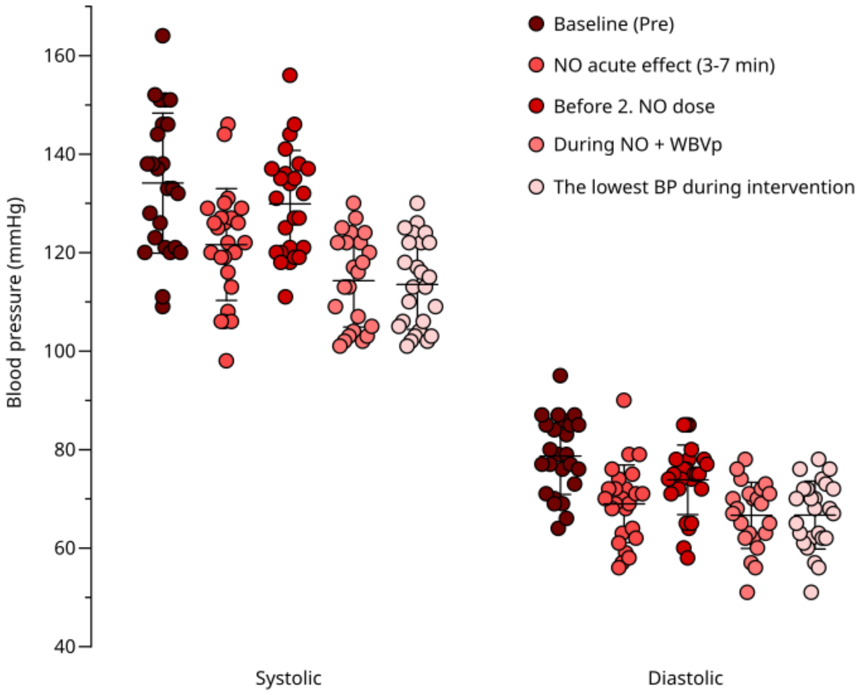


Figure S2. Measured blood pressure (systolic, diastolic) during NO+WBVp session.

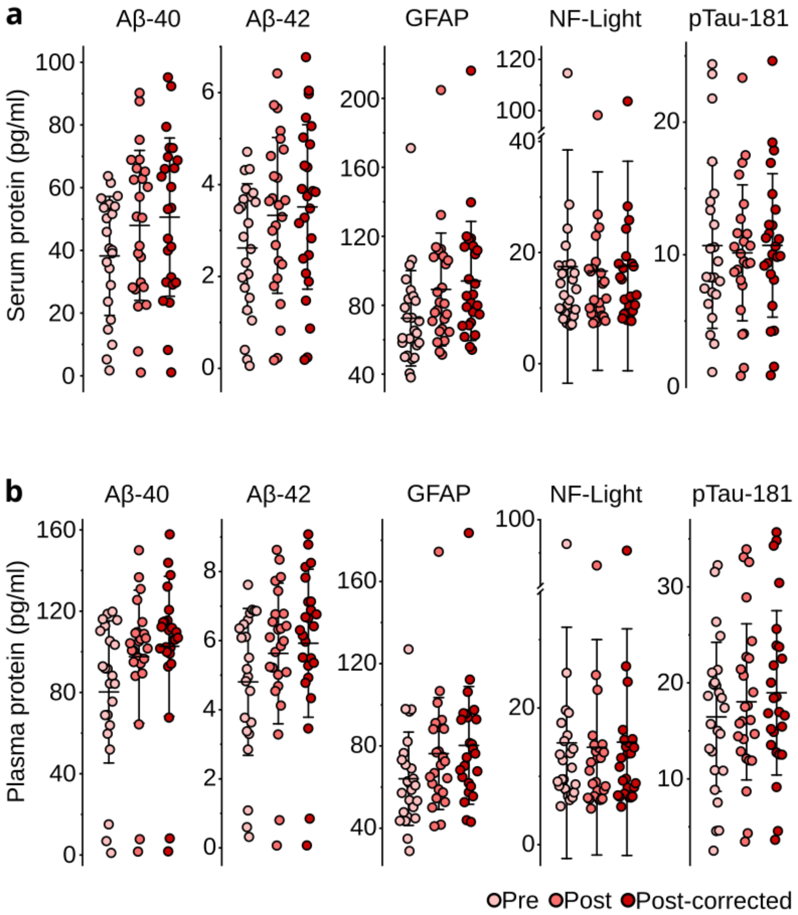


Figure S3. a. Serum and b. plasma concentrations of brain-derived proteins before (Pre) and after (Post) NO+WBVp intervention, with post-intervention values additionally shown after hemodilution correction.

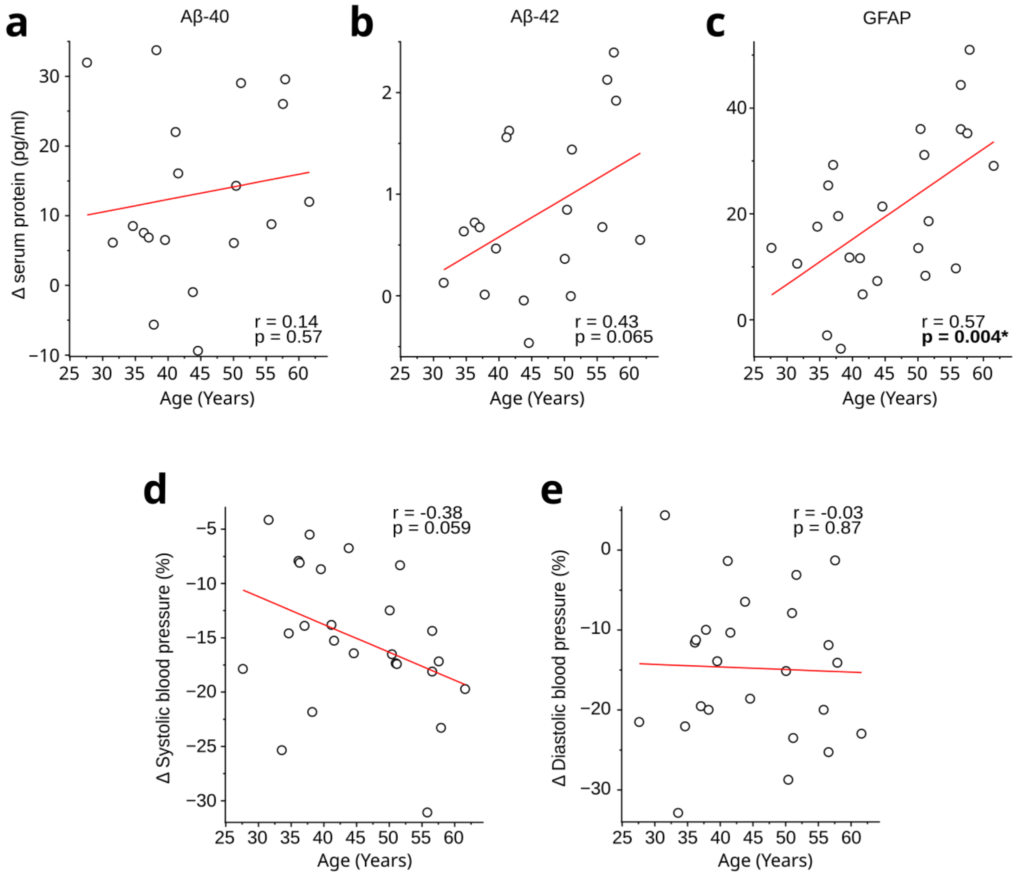


Figure S4. a-c. Changes in serum beta-amyloid (Aβ40, Aβ42) and glial fibrillary acidic protein (GFAP) as functions of age (Pearson's correlation) in NO+WBVp intervention. d-e. Changes in maximal reduction of systolic and diastolic blood pressure during NO+WBVp intervention as functions of age (Pearson's correlation).

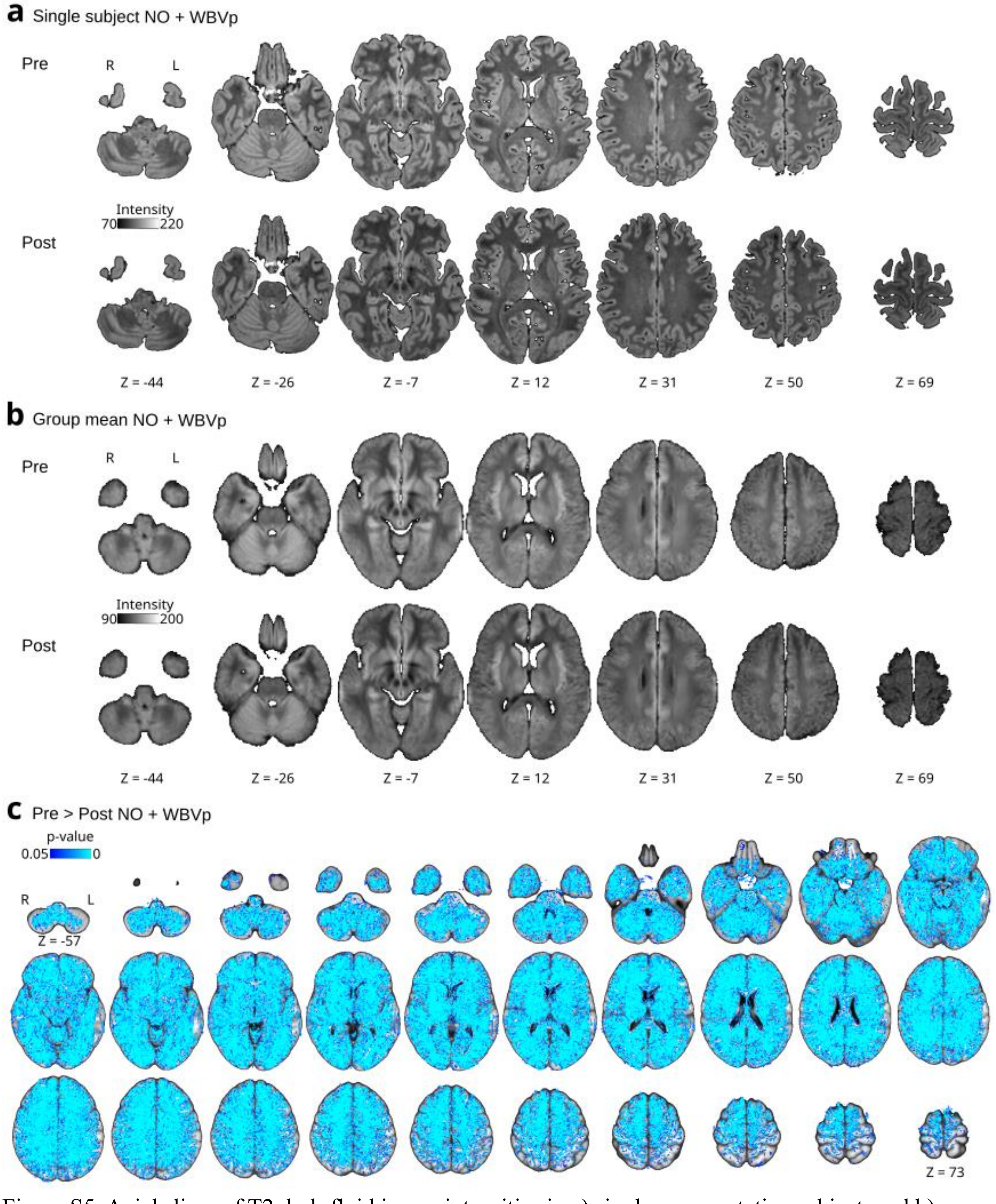


Figure S5. Axial slices of T2 dark-fluid image intensities in a) single representative subject, and b) group mean average (n = 24) T2DF images before (Pre) and after (Post) the combined vasomechanic NO+WBVp intervention. c) Statistical analysis result maps of the Post vs. Pre T2DF intensity maps (two-sample paired t-test, family-wise-error-rate correction (FWER), $p < 0.05$) indicating a significant whole brain level signal change from the intervention. R = right, L = left, Z = axial plane MNI152 coordinate.

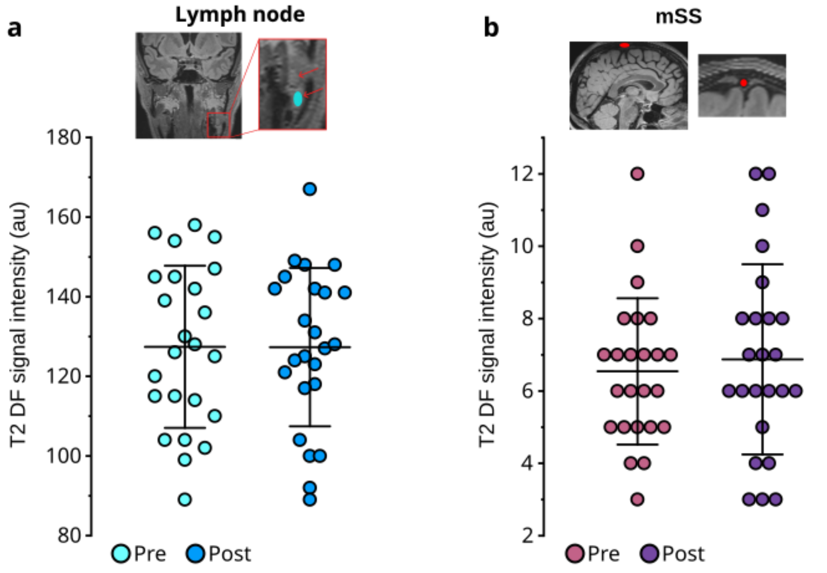


Figure S6. ROI-analysis of T2 dark-fluid images of a. cervical lymph node and b. middle/vertex superior sagittal sinus (mSS) before (Pre) and after (Post) NO+WBVp intervention.

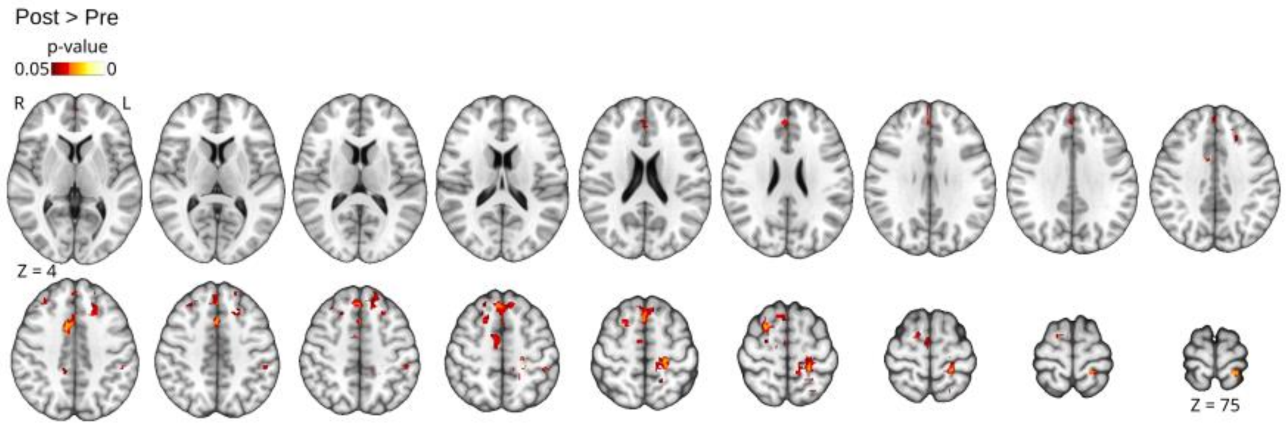


Figure S7. Spatial distribution of the cardiac pulsation brain power difference maps of NO+WBVp (n = 23) between Pre and Post intervention. R = right, L = left, Z = axial plane MNI152 coordinate.

**Nitric oxide (NO) mediated vasodilation**

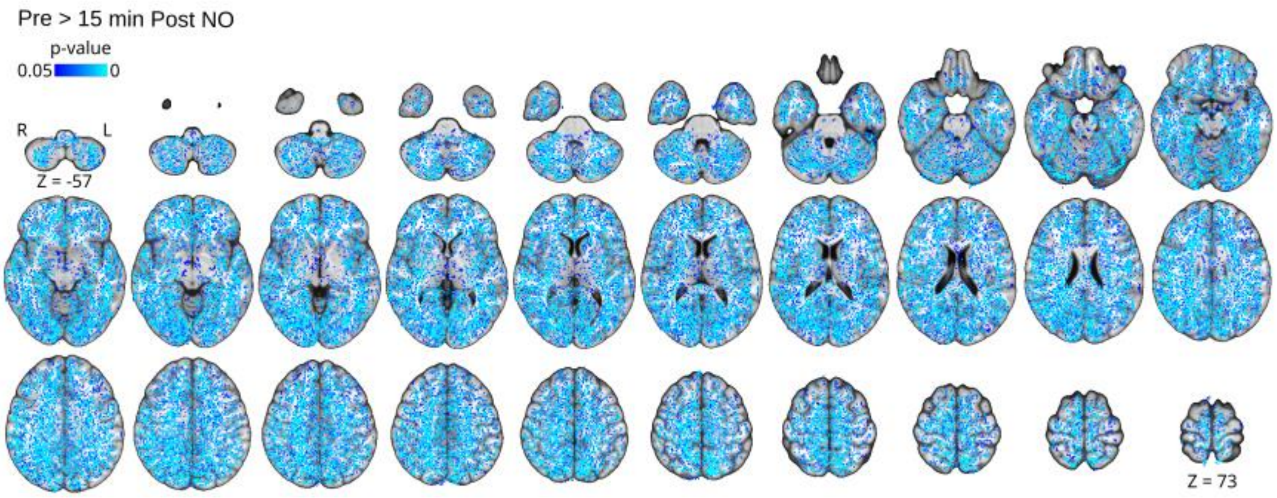


Figure S8. Spatial distribution of T2 dark-fluid difference maps of NO (n = 11) intervention between Pre and 15 min Post NO administration. R = right, L = left, Z = axial plane MNI152 coordinate.

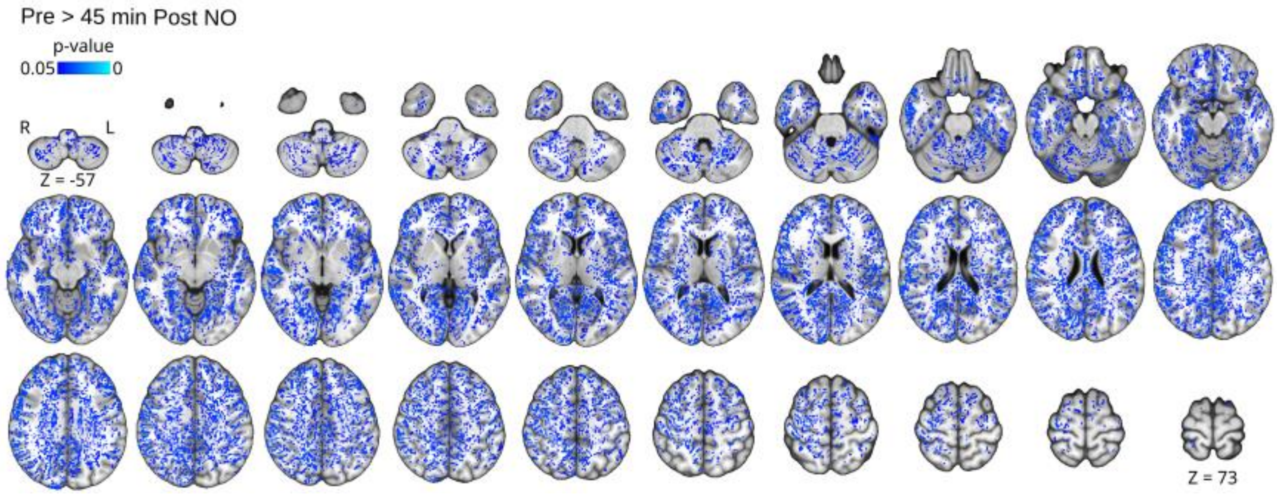


Figure S9. Spatial distribution of T2 dark-fluid difference maps of NO (n = 12) intervention between Pre and 45 min Post NO administration. R = right, L = left, Z = axial plane MNI152 coordinate.

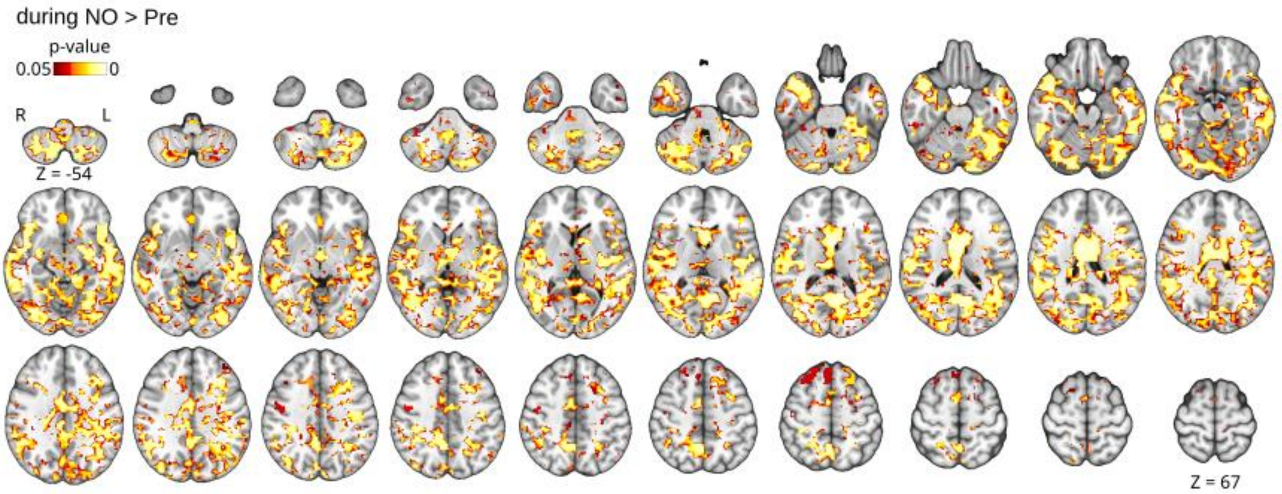


Figure S10. Spatial distribution of the low frequency (LF) brain pulsation (0.08-0.15 Hz) power difference maps for the NO intervention (n = 19) between Pre and during NO conditions. R = right, L = left, Z = axial plane MNI152 coordinate.

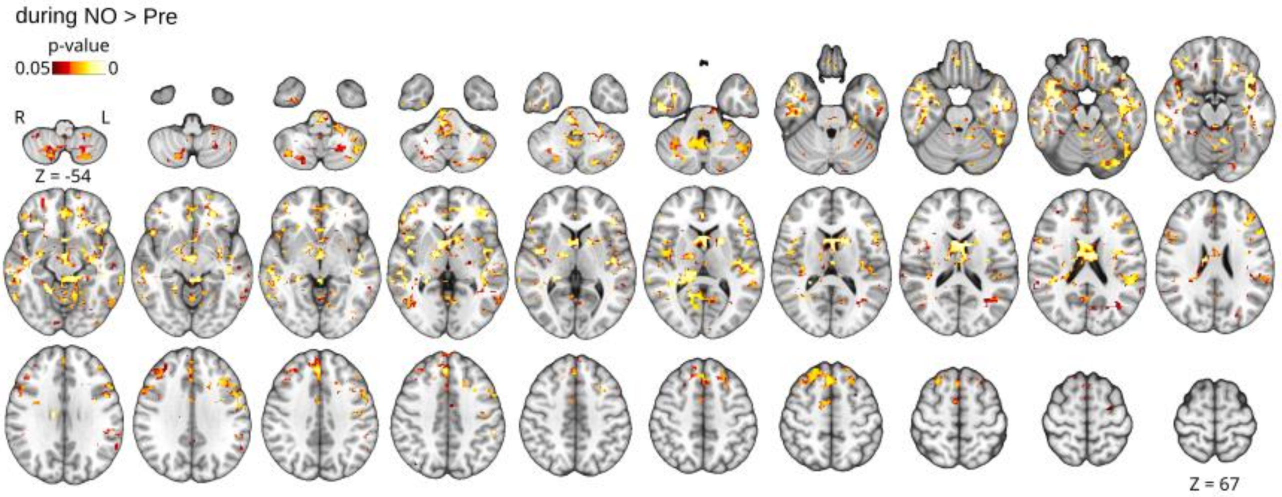


Figure S11. Spatial distribution of the respiratory pulsation brain power difference maps of the NO intervention (n = 19) between Pre and during NO conditions. R = right, L = left, Z = axial plane MNI152 coordinate.

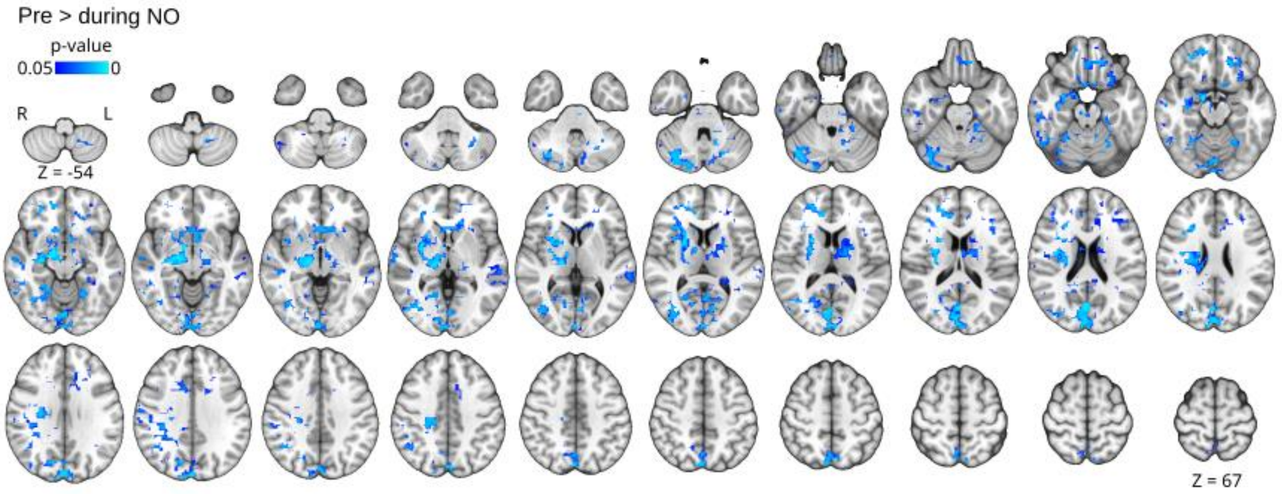


Figure S12. Spatial distribution of the cardiac pulsation brain power difference maps of the NO intervention (n = 23) between Pre and during the NO condition. R = right, L = left, Z = axial plane MNI152 coordinate.

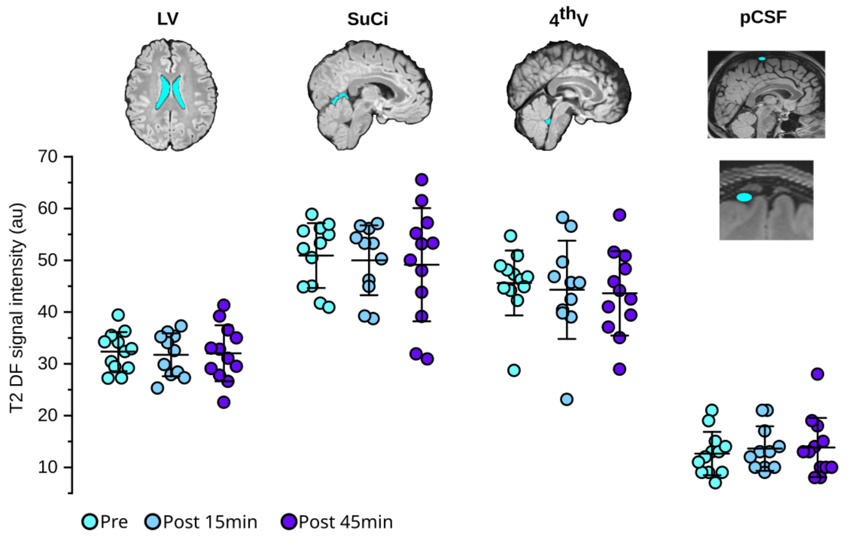


Figure S13. Region-of-interest analysis of T2 dark-fluid images of lateral ventricles (LV), superior cisterna (SuCi), 4th ventricle (4thV), and middle parasagittal CSF space (pCSF) before (Pre) and after NO administration at 15 min (Post 15 min) and 45 min (Post 45 min).

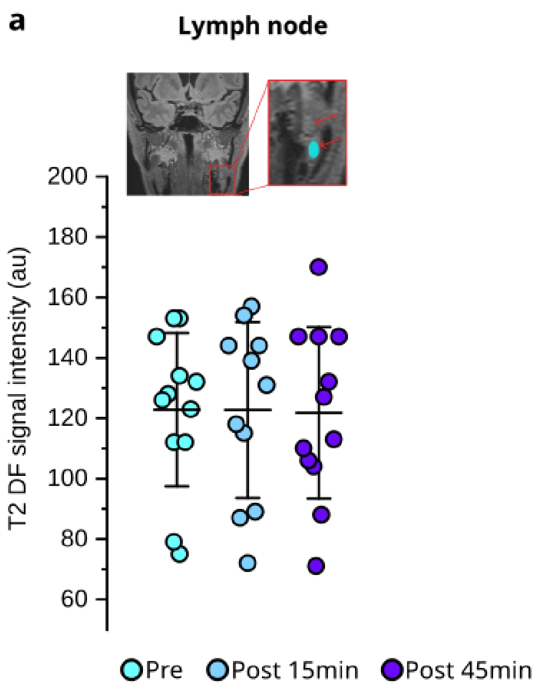


Figure S14. Region-of-interest analysis of T2 dark-fluid images of a. superficial cervical lymph nodes and b. para-cavernous dura CSF-filled spaces (PCD) before (Pre) and after NO administration at 15 min (Post 15 min) and 45 min (Post 45 min).

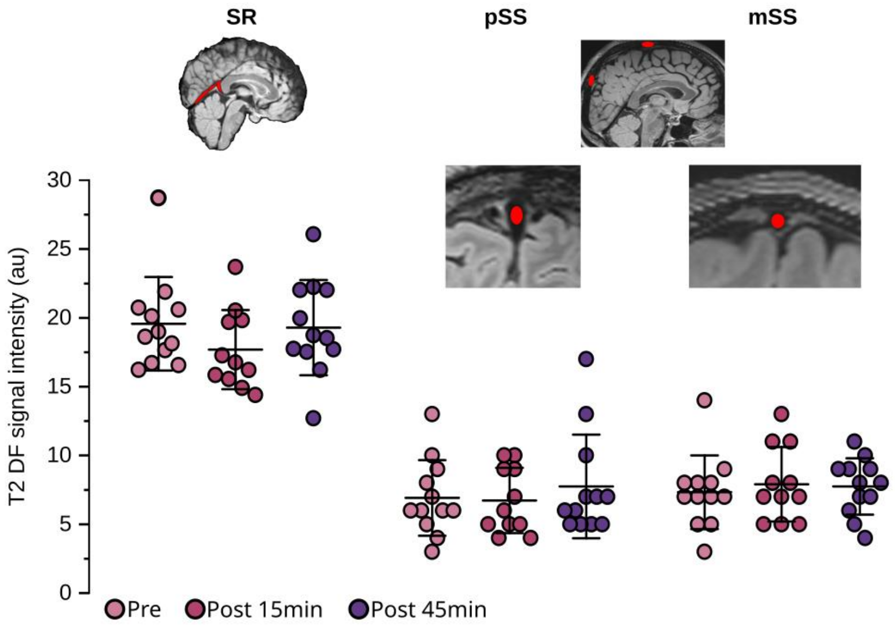


Figure S15. Region-of-interest analysis of T2 dark-fluid images of sinus rectus (SR), posterior sagittal sinus (pSS), and middle/vertex superior sagittal sinus (mSS) before (Pre) and after NO administration at 15 min (Post 15 min) and 45 min (Post 45 min). As the blood underwent dilution based on the laboratory findings (c.f. Fig 3l), the diluted blood should have been darker. However, the lack of expected T2DF signal darkening suggests that there was an increased protein content in the blood, which compensated for the increase of free water in the blood, a conjecture which was also verified with the Simoa laboratory findings (Fig 1d.). Thus, the net darkening of the whole brain cannot be due to increased brain tissue blood, but likely reflects the reduced amount of brain water/protein in the NO treatment condition.

**Piezo-1 targeted whole body vibration (WBVp)**

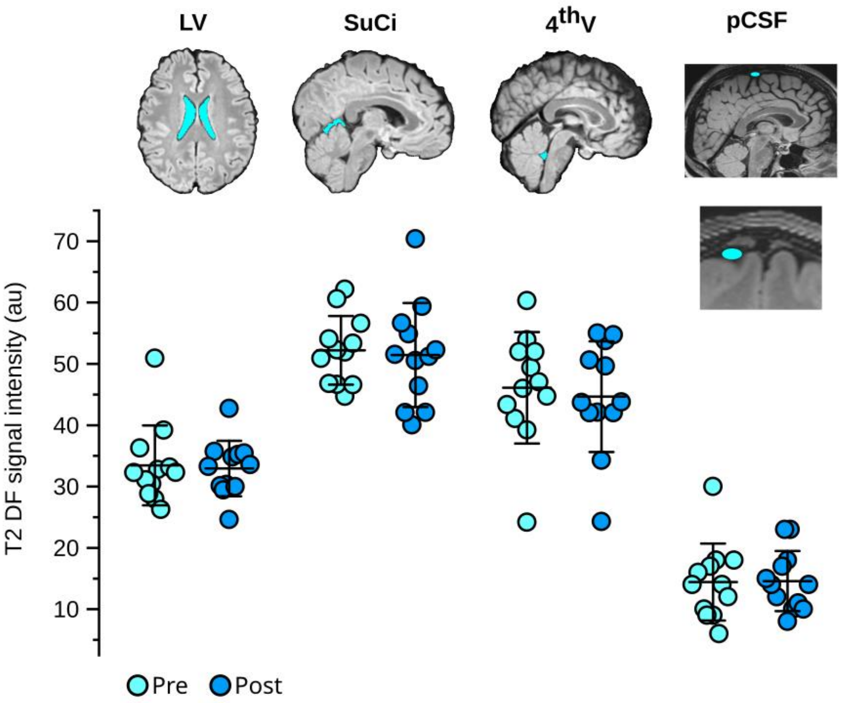


Figure S16. Region-of-interest analysis of T2 dark-fluid images of lateral ventricles (LV), superior cisterna (SuCi), 4th ventricle (4thV), and middle parasagittal CSF space (pCSF) before (Pre) and after (Post) Piezo-1 targeted whole body vibration intervention.

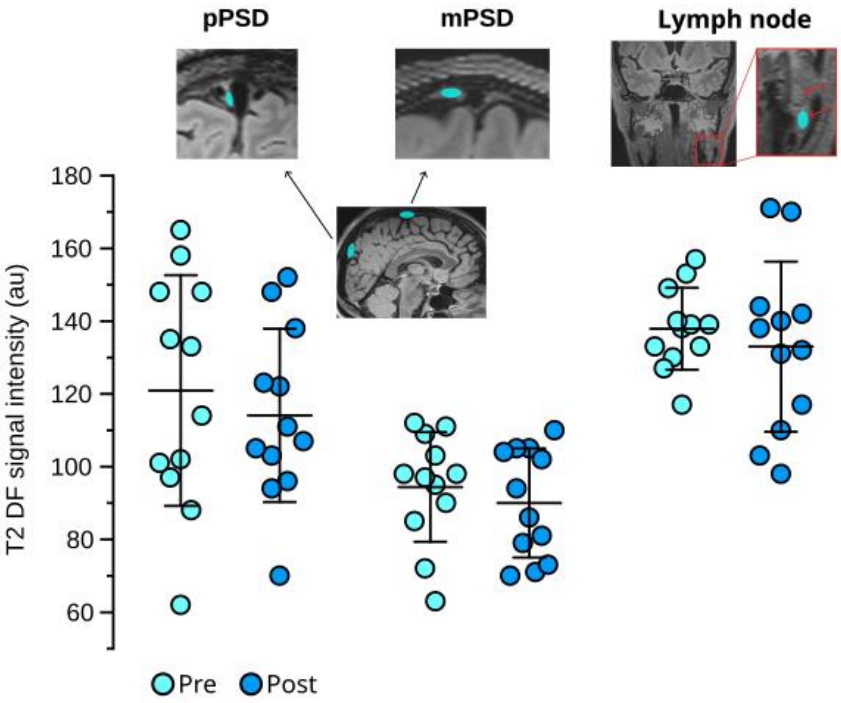


Figure S17. Region-of-interest analysis of T2 dark-fluid images of posterior parasagittal dura CSF-filled space (pPSD), middle/vertex parasagittal dura CSF-filled space (mPSD), and superficial cervical lymph nodes before (Pre) and after (Post) Piezo-1 targeted whole body vibration intervention.

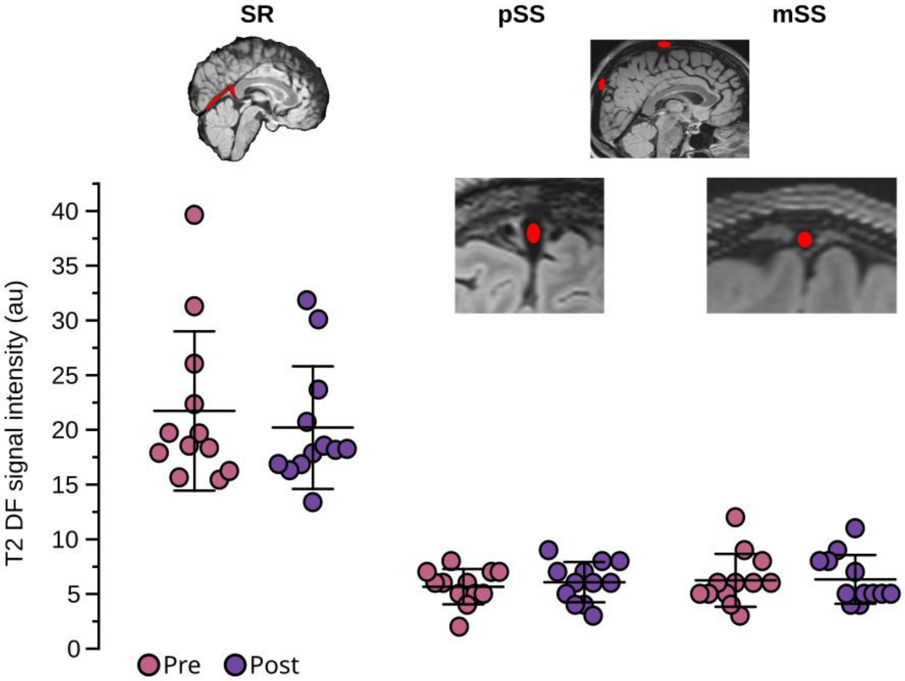

Figure S18. Region-of-interest analysis of T2 dark-fluid images of sinus rectus (SR), posterior sagittal sinus (pSS) and middle/vertex superior sagittal sinus (mSS) before (Pre) and after (Post) Piezo-1 targeted whole body vibration intervention.

**Material and methods**

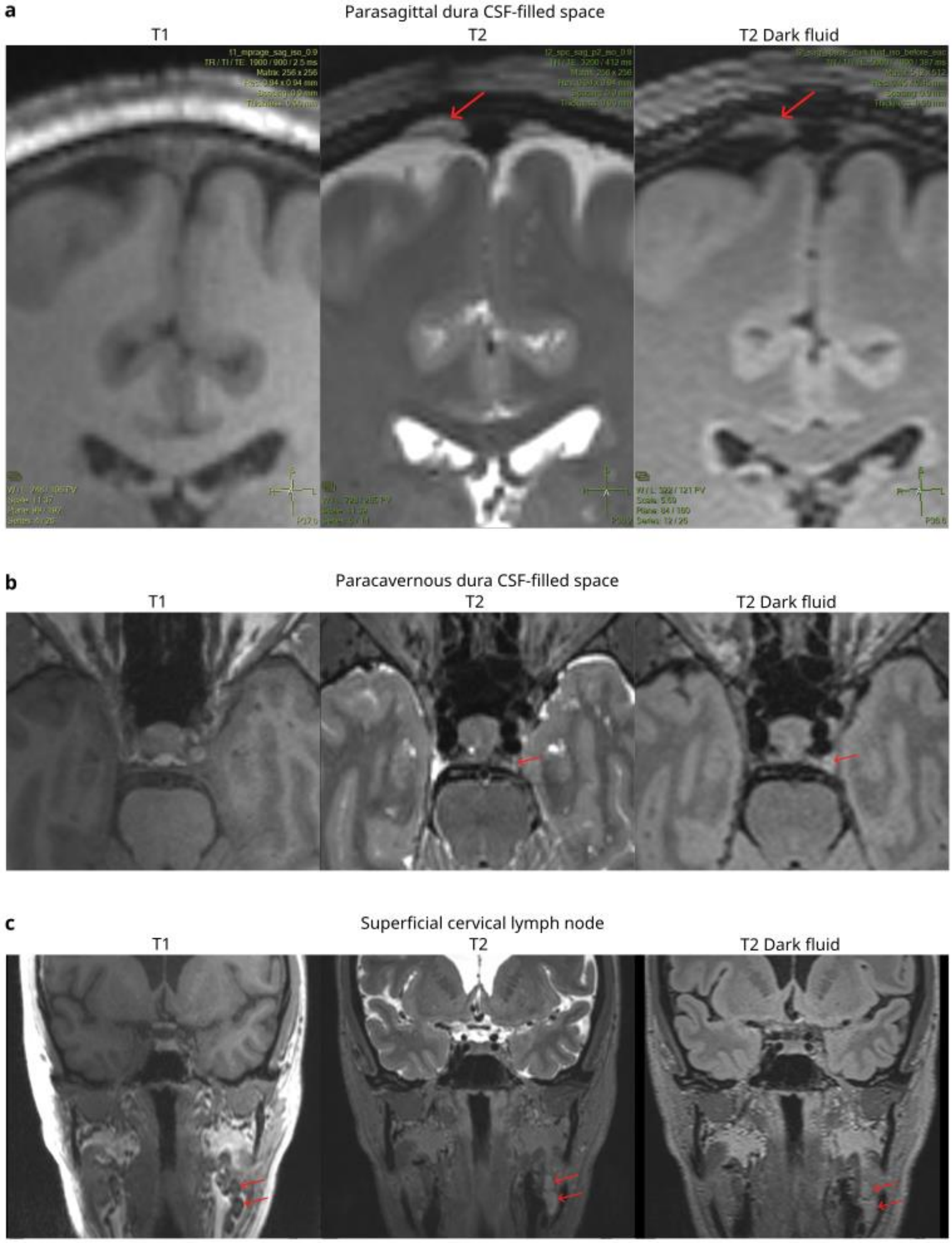


Figure S19. Representative images to define a. parasagittal dura CSF-filled space, b. para-cavernous dura CSF-filled space, and c. superficial cervical lymph nodes from T2 dark-fluid images using T1- and T2-weighted images.

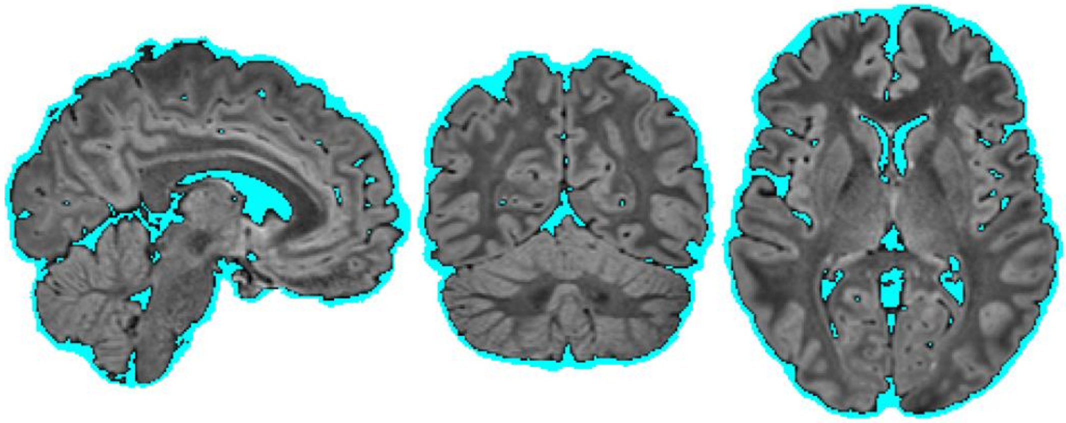

Figure S20. Representative segmented brain tissue (grey) and CSF (light blue) from one participant.

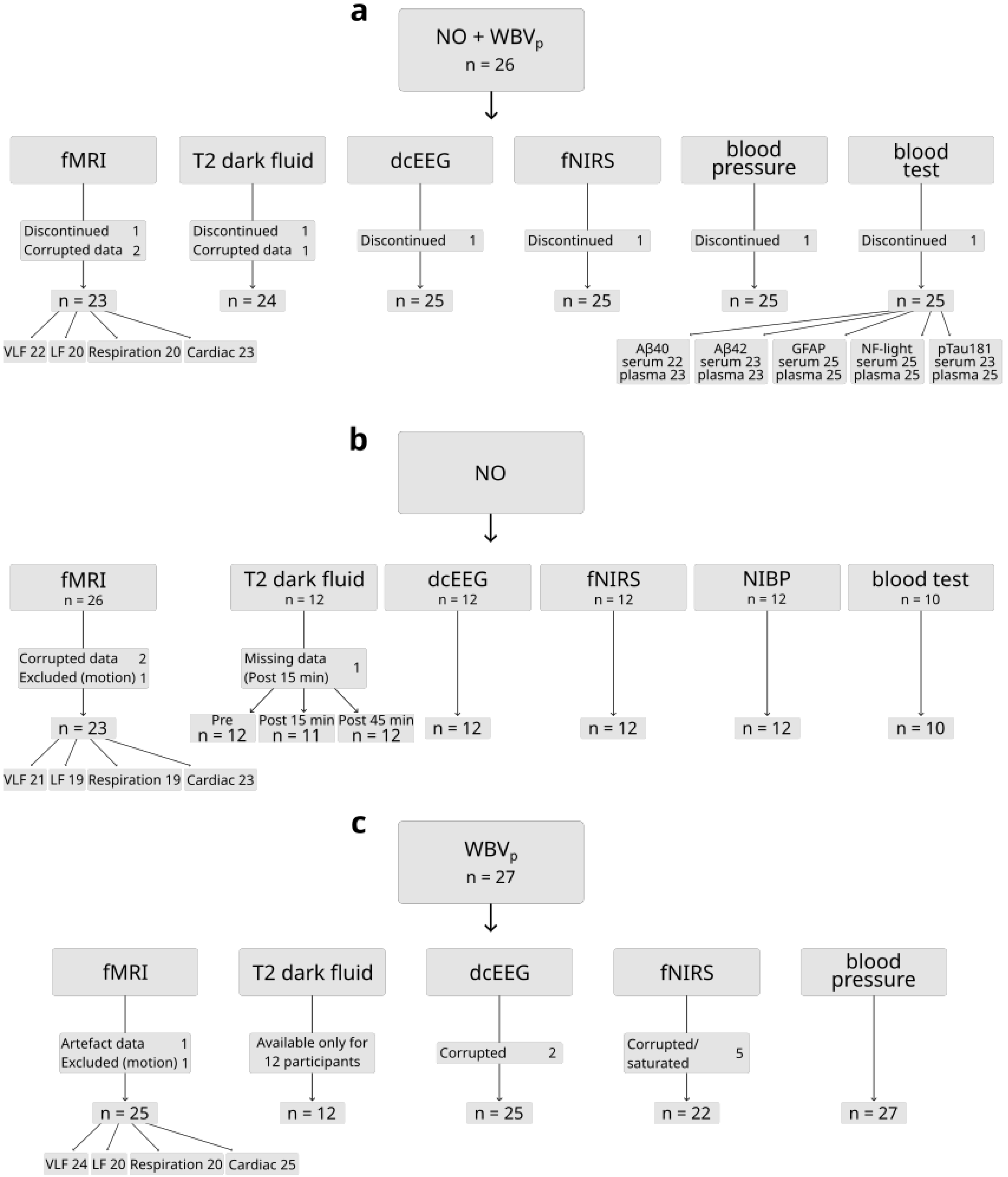


Figure S21. Available multimodal data samples for analysis and statistics of **a.** NO-mediated vasodilation combined to Piezo-1 targeted whole-body vibration (NO+WBVp) intervention, **b.** NO-mediated vasodilation (NO) intervention, and **c.** Piezo-1 targeted whole-body vibration (WBVp) intervention.

**Mouse study**

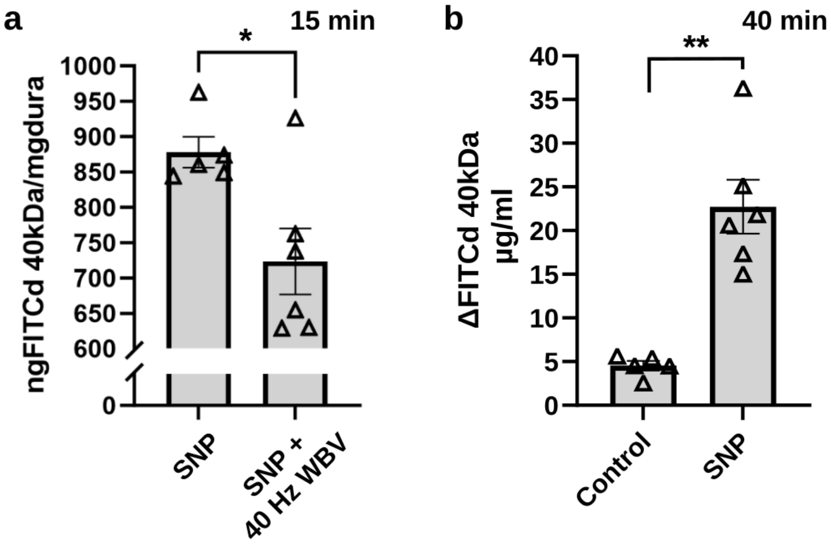


Figure S22. a. Vasodilation with sodium nitroprusside (SNP) combined with 40 Hz vibration treatment for 15 min results in significantly less tracer (FITC-dextran 40 kDA) accumulation in dorsal dura mater compared to mice treated with SNP alone. b. 40 kDa FITC-dextran concentration increased in blood during 40-min NO-mediated vasodilation with SNP compared to mice treated with saline. Unpaired two-tailed t-test with Welch's correction, * $p < 0.05$, ** $p< 0.01$, data are mean ± standard error of mean, n = 5-6 mice per group.
